# The best COVID-19 predictor is recent smell loss: a cross-sectional study

**DOI:** 10.1101/2020.07.22.20157263

**Authors:** Richard C. Gerkin, Kathrin Ohla, Maria G. Veldhuizen, Paule V. Joseph, Christine E. Kelly, Alyssa J. Bakke, Kimberley E. Steele, Michael C. Farruggia, Robert Pellegrino, Marta Y. Pepino, Cédric Bouysset, Graciela M. Soler, Veronica Pereda-Loth, Michele Dibattista, Keiland W. Cooper, Ilja Croijmans, Antonella Di Pizio, M. Hakan Ozdener, Alexander W. Fjaeldstad, Cailu Lin, Mari A. Sandell, Preet B. Singh, V. Evelyn Brindha, Shannon B. Olsson, Luis R. Saraiva, Gaurav Ahuja, Mohammed K. Alwashahi, Surabhi Bhutani, Anna D’Errico, Marco A. Fornazieri, Jérôme Golebiowski, Liang-Dar Hwang, Lina Öztürk, Eugeni Roura, Sara Spinelli, Katherine L. Whitcroft, Farhoud Faraji, Florian Ph.S Fischmeister, Thomas Heinbockel, Julien W. Hsieh, Caroline Huart, Iordanis Konstantinidis, Anna Menini, Gabriella Morini, Jonas K. Olofsson, Carl M. Philpott, Denis Pierron, Vonnie D.C. Shields, Vera V. Voznessenskaya, Javier Albayay, Aytug Altundag, Moustafa Bensafi, María Adelaida Bock, Orietta Calcinoni, William Fredborg, Christophe Laudamiel, Juyun Lim, Johan N. Lundström, Alberto Macchi, Pablo Meyer, Shima T. Moein, Enrique Santamaría, Debarka Sengupta, Paloma Rohlfs Dominguez, Hüseyin Yanik, GCCR Group Author, Thomas Hummel, John E. Hayes, Danielle R. Reed, Masha Y. Niv, Steven D. Munger, Valentina Parma, Non-byline authors (to be listed as collaborators in PubMed under the GCCR Group Author)

**Affiliations:** School of Life Sciences, Arizona State University; Institute of Neuroscience and Medicine (INM3), Forschungszentrum Jülich; Department of Anatomy, Mersin University; National Institutes of Nursing Research; National Institute of Alcohol Abuse and Alcoholism; National Institutes of Health; AbScent; Food Science, The Pennsylvania State University; National Institute of Diabetes and Digestive and Kidney Diseases; Interdepartmental Neuroscience Program, Yale University; Food Science, University of Tennessee; Department of Food Science and Human Nutrition, University of Illinois at Urbana Champaign; Institut de Chimie de Nice, UMR CNRS 7272, Université Côte d’Azur; Grupo de Estudio de Olfato y Gusto (GEOG), Buenos Aires; Department of Otorhinolaryngology University of Buenos Aires; Medecine Evolutive UMR5288, University of Toulouse; Department of Basic Medical Science, Neuroscience and Sense Organs, Università degli Studi di Bari A. Moro; Department of Neurobiology and Behavior, University of California, Irvine; Department of Psychology, Utrecht University; Leibniz-Institute for Food Systems Biology at the Technical University of Munich; Monell Chemical Senses Center; Flavour Clinic, Department of Otorhinolaryngology, Regional Hospital West Jutland; Department of Food and Nutrition, University of Helsinki; Department of Oral Surgery and Oral Medicine, Faculty of Dentistry, University of Oslo; Department of Electrical and Electronics Engineering, Karunya Institute of Technology and Sciences; National Centre for Biological Sciences, Tata Institute of Fundamental Research; Research Branch, Sidra Medicine; Computational Biology, Indraprastha Institute of Information Technology; ENT Division, Surgery Department, Sultan Qaboos University; School of Exercise and Nutritional Sciences, San Diego State University; Cellular and Molecular Neurobiology, Goethe University Frankfurt; Clinical Surgery, Universidade Estadual de Londrina; The University of Queensland Diamantina Institute, The University of Queensland; Centre for Nutrition and Food Sciences, Queensland Alliance for Agriculture and Food Innovation, The University of Queensland; Department of Agriculture, Food, Environment and Forestry (DAGRI), University of Florence; Ear Institute, University College London; Department of Surgery, Division of Otolaryngology-Head and Neck Surgery, UC San Diego Health; Department of Psychology, University of Graz; Department of Anatomy, Howard University College of Medicine; Department of Otorhinolaryngology, Rhinology-Olfactology Unit, Geneva University Hospitals; ENT Department, Cliniques universitaires Saint-Luc; ORL Department, Papageorgiou Hospital, Aristotle University; Neuroscience Area, SISSA, International School for Advanced Studies; University of Gastronomic Sciences; Department of Psychology, Stockholm University; Norwich Medical School, The Norfolk Smell & Taste Clinic, University of East Anglia; Biological Sciences Department, Fisher College of Science and Mathematics, Towson University; Severtsov Institute of Ecology and Evolution RAS; Department of General Psychology, University of Padova; Otorhinolaryngology Department, Biruni University; Lyon Neuroscience Research Center, CNRS; Departamento de Salud Pública ORL, Hospital General Barrio Obrero; Private practice, Milan; Scent Engineering, DreamAir LLC; Food Science and Technology, Oregon State University; Department of Clinical Neuroscience, Karolinska Institutet; ENT Department University of Insubria Varese, Assi-Sette Laghi; Italian Academy of Rhinology; Health Care and Life Sciences, IBM TJ. Watson Research Center; School of Biological Sciences, Institute for Research in Fundamental Sciences; Clinical Neuroproteomics Unit, Navarrabiomed-IdiSNA; Psychology and Anthropology, University of Extremadura; Department of Electrical and Electronics Engineering, Mersin University; Division of Human Nutrition and Health, Wageningen University; Behavioral Science Institute, Radboud University; Department of Psychiatry and Psychotherapy, Friedrich-Alexander- Universität Erlangen-Nürnberg; Centro de Otorrinolaringología Respira Libre; Department of Biology, University of Chile; Department of Physiology, The Federal University of Technology, Akure, Nigeria; CSGA-Centre for Taste and Feeding Behavior, INRAE, CNRS, AgroSup Dijon, University Bourgogne Franche- Comté; Department of Psychology, University of Dayton; ENT and Head and Neck Research Center and Department, the Five Senses Institute, Iran University of Medical Sciences; Neuroscience Department, Federico II University; Department of Neuroscience, Biomedicine and Movement Sciences, Anatomy and Histology Section, University of Verona; Department of Otolaryngology-Head and Neck Surgery, Second Affiliated Hospital of Xi’an Jiaotong University; Food Technology, IRTA; Department of Anthropology, University of Alaska Fairbanks; Department of Medicine, Hadassah - Hebrew University Medical Center, Mt Scopus; University of Massachusetts; Servicio de Otorrinolaringología, Hospital Italiano de Buenos Aires; Department of Otolaryngology-Head and Neck Surgery, Johns Hopkins University School of Medicine; School of Cognitive Sciences, Institute for Research in Fundamental Sciences (IPM); Agriculture and Food, Commonwealth Scientific and Industrial Research Organisation (CSIRO); Department of Psychiatry, The Affiliated Brain Hospital of Guangzhou Medical University; Center for Smell and Taste, University of Florida; Institute of Food and Health, University College Dublin; Department of Anatomy, Université du Québec à Trois-Rivières; ENT Department, Guy’s and St Thomas’ Hospitals; Department of Biochemistry, Food Science and Nutrition, The Hebrew University of Jerusalem; Department of Otorhinolaryngology, Smell and Taste Clinic, TU Dresden; Smell & Taste Clinic, Department of Otorhinolaryngology, TU Dresden; Department of Molecular Medicine, University of Padova; Department of Neurology, The Affiliated Brain Hospital of Guangzhou Medical University; Nutrition and Dietetics, Kilis 7 Aralik University; Sensory Neuroscience Laboratory, University of Otago; Department of Otorhinolaryngology, Sancaktepe Education and Research Hospital; University of Pennsylvania, Perelman School of Medicine; CMOENES groups, Centre de Recherche en Neurosciences de Lyon; Department of Otolaryngology, University of Florida; Department of Chemistry, Indiana University; Department of Otolaryngology - Head & Neck Surgery, Columbia University Medical Center; Centre for Global Health, Usher Institute, University of Edinburgh; Department of Public Health, University of California Merced; Division of Psychology, University of Stirling; Centre for the Study of the Senses, University of London; Department of Statistics, Florida State University; First Department of Specialties, Hôpital Provincial Général de Bukavu, Faculty of Medicine, Université Catholique de Bukavu; The Faculty of Agriculture, Food and Environment, The Hebrew University of Jerusalem; Department of Pharmacology and Therapeutics, University of Florida, Department of Psychology; Temple University

## Abstract

**Background:** COVID-19 has heterogeneous manifestations, though one of the most common symptoms is a sudden loss of smell (anosmia or hyposmia). We investigated whether olfactory loss is a reliable predictor of COVID-19.

**Methods:** This preregistered, cross-sectional study used a crowdsourced questionnaire in 23 languages to assess symptoms in individuals self-reporting recent respiratory illness. We quantified changes in chemosensory abilities during the course of the respiratory illness using 0-100 visual analog scales (VAS) for participants reporting a positive (C19+; n=4148) or negative (C19-; n=546) COVID-19 laboratory test outcome. Logistic regression models identified singular and cumulative predictors of COVID-19 status and post-COVID-19 olfactory recovery.

**Results:** Both C19+ and C19-groups exhibited smell loss, but it was significantly larger in C19+ participants (mean±SD, C19+: -82.5±27.2 points; C19-: -59.8±37.7). Smell loss during illness was the best predictor of COVID-19 in both single and cumulative feature models (ROC AUC=0.72), with additional features providing negligible model improvement. VAS ratings of smell loss were more predictive than binary chemosensory yes/no-questions or other cardinal symptoms, such as fever or cough. Olfactory recovery within 40 days was reported for ∼50% of participants and was best predicted by time since illness onset.

**Conclusions:** As smell loss is the best predictor of COVID-19, we developed the ODoR-19 tool, a 0-10 scale to screen for recent olfactory loss. Numeric ratings ≤2 indicate high odds of symptomatic COVID-19 (4<OR<10), which can be deployed when viral lab tests are impractical or unavailable.

## Introduction

The novel coronavirus SARS-CoV-2 responsible for the global COVID-19 pandemic has left a staggering level of morbidity, mortality, and societal and economic disruption in its wake.^1^ Early publications^2–8^ indicate that sudden smell and taste loss are cardinal, early and potentially specific symptoms of COVID-19,^9^ including in otherwise asymptomatic individuals.^10–13^ While fever and cough are common symptoms of diverse viral infections, the potential specificity of early chemosensory loss to COVID-19 could make it valuable in screening and diagnosis.

Anosmia and other chemosensory disorders have serious health and quality-of-life consequences for patients. However, the general lack of awareness of anosmia and other chemosensory disorders by clinicians and the public, including their association with upper respiratory infections,^14^ contributed to an underappreciated role of chemosensory symptoms in the diagnosis of COVID-19. Additionally, the impact of smell loss as a clinical consequence of COVID-19 has not been adequately addressed. Thus, there is an urgent need to better define the chemosensory dysfunctions associated with COVID-19 and to determine their relevance as predictors of this disease. It is critical to develop rapid clinical tools to efficiently and effectively integrate chemosensory assessments into COVID-19 screening and treatment protocols. Information on the duration and reversibility of post-COVID-19 chemosensory impairment is also lacking.

We used binary, categorical and continuous self-report measures to determine the chemosensory phenotype, along with other symptoms and characteristics, of COVID-19-positive (C19+) and COVID-19-negative (C19-) individuals who had reported recent symptoms of respiratory illness. Using those results in logistic regression models, we identified predictors of COVID-19 and recovery from smell loss. Finally, we propose the **O**lfactory **D**eterminati**o**n **R**ating scale for COVID-19 (ODoR-19), a quick, simple-to-use, telemedicine-friendly tool to improve the utility of current COVID-19 screening protocols, particularly when access to rapid testing for SARS-CoV-2 is limited.

## Methods

### Study design

This preregistered,^15^ cross-sectional online study was approved by the Office of Research Protections of The Pennsylvania State University (STUDY00014904); it is in accordance with the revised Declaration of Helsinki, and compliant with privacy laws in the U.S.A. and European Union. Data reported here were collected between April 7 and July 3, 2020 from the Global Consortium for Chemosensory Research (GCCR) core questionnaire (**Appendix 1** and https://gcchemosensr.org),^8^ an online crowdsourced survey deployed in 32 languages that used binary response and categorical questions (e.g. **Appendix 1**, Questions 6,9) and visual analog scales (e.g., **Appendix 1**, Question 13) to measure self-reported chemosensory ability and other symptoms in adults with current or recent respiratory illness. Data reported here include responses in Arabic, Bengali, Chinese (Simplified and Traditional), Danish, Dutch, English, Farsi, Finnish, French, German, Greek, Hebrew, Hindi, Italian, Japanese, Korean, Norwegian, Portuguese, Russian, Spanish, Swedish, Turkish, and Urdu.

### Participants (272)

A convenience sample of 52,334 volunteers accessed the GCCR questionnaire; 25,620 met criteria (≥19 years old, respiratory illness or suspicion thereof within the past two weeks). After applying pre-registered exclusion criteria, 15,747 participants were included in reported analyses (**Figure 1**).

**Figure 1.**
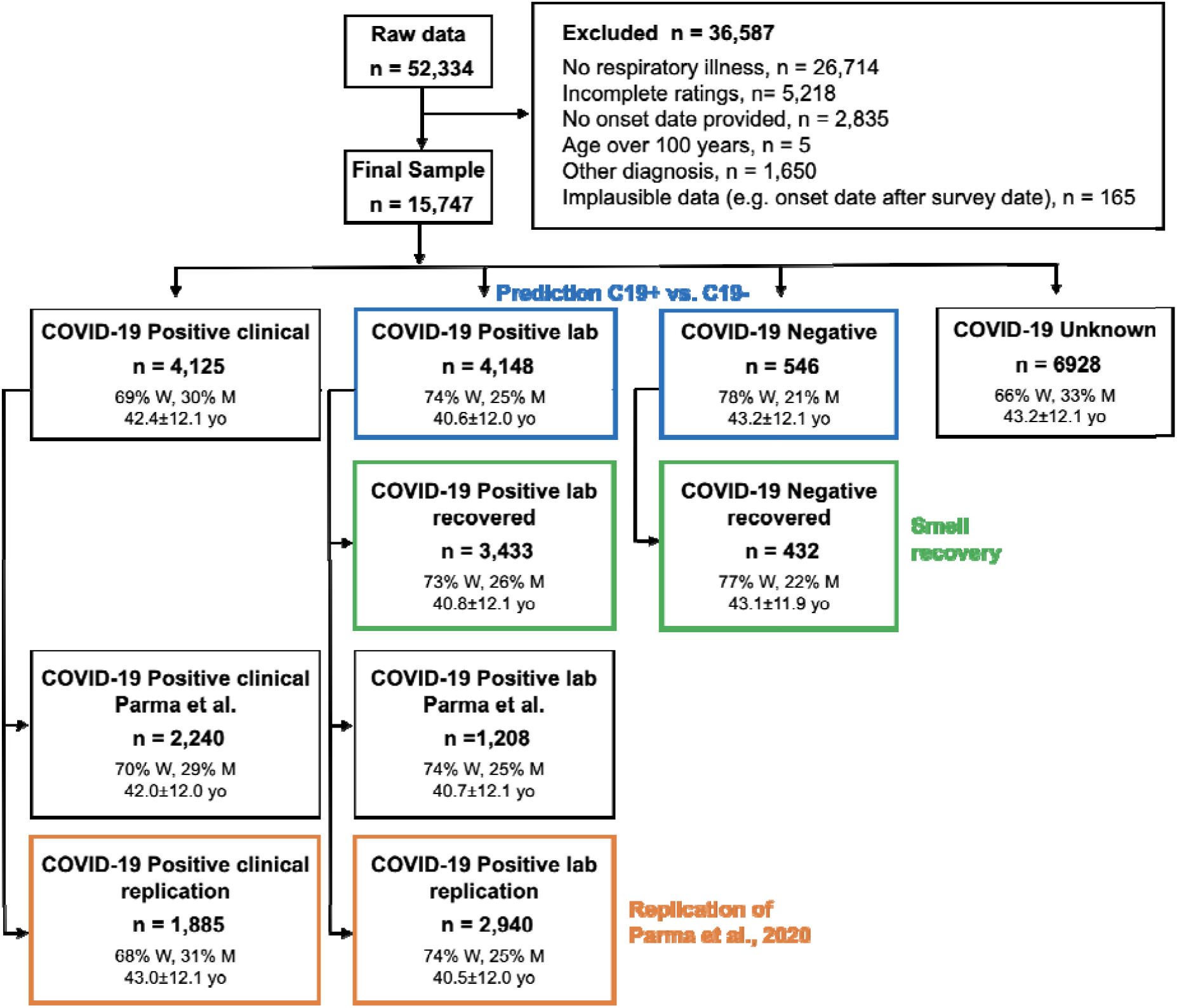
Flow diagram showing the demographics of participants included and excluded in the present analyses. Participants included in the prediction of COVID-19 status are highlighted in blue. Participants included in the smell recovery models are highlighted in green. Participants included in the replication of our prior work are highlighted in orange. N = number of participants; yo = age in years; W = women; M = men. Gender percentages do not include <1% of participants who answered “other” or “preferred not to say”. Participants described in the green boxes are a subset of those described in the boxes listed above them.

Based on the self-reported outcome of a COVID-19 lab test, participants were labeled as either C19+ (positive result) or C19- (negative result). The specific collider bias characterizing this sample (high fraction of C19+ participants and high prevalence of chemosensory disorders in both groups) underestimates the positive correlation between smell loss and COVID-19 (**Figure S1**). Thus, it represents a conservative scenario to test the hypothesis that smell loss reliably predicts COVID-19 status. We benchmarked the GCCR dataset to the representative samples collected with the Imperial College London YouGov Covid 19 Behaviour Tracker (henceforth, YouGov; countries shared across datasets: Brazil, Canada, Denmark, Finland, France, Germany, Italy, Mexico, Netherlands, Norway, Spain, Sweden, UK, USA; YouGov: N=8,674, GCCR: N=3,962; data publicly available at https://github.com/YouGov-Data/covid-19-tracker). Benchmarking shows the GCCR sample underestimates the positive association between smell loss and C19+ (**Figure S1, Table S1**). The country-wise fraction of C19+ participants is correlated (r∼0.45) when responses from the same calendar week are aligned (**Figure S2)**. These findings are in line with other comparisons between crowdsourced versus representative health data,^16^ confirming that trends identified in crowdsourced data reasonably approximate population data. Because the GCCR cohort is not demographically balanced, it should not be used to estimate prevalence. However, the representative YouGov cohort indicates globally ∼33% of C19+ individuals report smell loss (**Table S1**).

### Statistical analyses

Statistical analyses were performed in Python 3.7.6 using the pandas,^17^ scikit-learn,^18^ and statsmodels^19^ packages. The data and annotated code is included as supplemental material and will be publicly available on GitHub (http://github.com/GCCR/GCCR002) upon publication. Missing values for covariates used in prediction models of COVID-19 status and smell recovery were imputed as follows: binary features = 0.5, numeric features = median, categorical variables = “Missing”. Prediction targets themselves were never imputed. Responses incompatible with model generalization (e.g., open ended questions) were excluded. A one-hot encoding was applied to all categorical variables to produce binary indicators of category membership. L1-regularized logistic regression (penalty α=1) consistently produced sparse models with comparable cross-validation accuracy and were therefore the prediction test of choice. Model quality was measured using receiver operating characteristic (ROC) area under the curve (AUC). Cross-validation was performed in 100 random splits of 80% training set and 20% test set, and ROC curves are concatenated over each test set. ROC curves are computed on predicted probabilities from each model, circumventing the high-cardinality bias of AUC. For single feature models, AUC is independent of most modeling details, including all rank-invariant decisions. To correctly compute p-values for model coefficients, the normalized data were standardized (mean 0, variance 1) and then coefficients back-transformed to normalized form after fitting.

## Results

### Chemosensory loss associates with COVID-19

A preregistered replication of our prior study^8^ confirmed that reported smell, taste, and chemesthesis abilities drop significantly in both lab-tested C19+ participants and those diagnosed by clinical assessment (**Figure S3, Table S2**).

Next, we compared lab-tested C19+ and C19- participants. C19+ participants reported a greater loss of smell (C19+: -82.5±27.2 points; C19-: -59.8±37.7 points; p=2.2e-46, extreme evidence of difference: BF_10_=8.97e+61; **Figure 2A,B; Table S3**), taste (C19+: -71.6±31.8 points; C19-: -55.2±37.5 points; p=6.7e-26, extreme evidence of difference: BF_10_=6.67e+24; **Figure 2C,D; Table S3**) and chemesthesis ability (C19+: - 36.8±37.1 points; C19-: -28.7±37.1 points; p=1.6e-07, extreme evidence of difference: BF_10_=3182; **Figure 2E,F; Table S3**). However, both groups reported a similar degree of nasal obstruction (**Figure 2G,H; Table S3**). Self-reported changes in smell, taste, and chemesthesis were highly correlated within both groups (C19+: 0.71<r<0.83; C19-: 0.76<r<0.87) and orthogonal to nasal obstruction changes (C19+: r=-0.20; C19-: r=-0.13).

**Figure 2.**
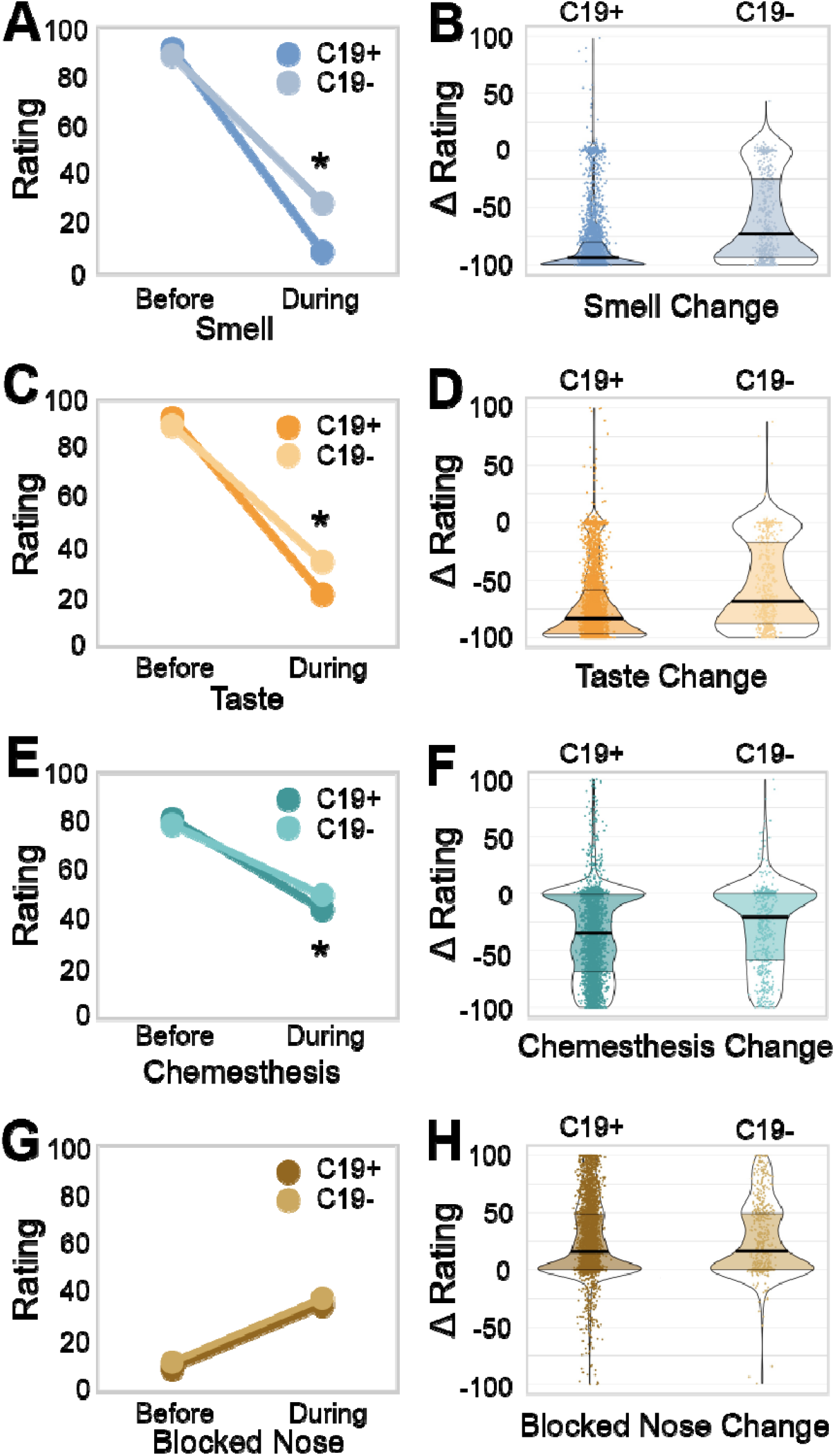
Chemosensory ability and nasal obstruction in C19+ and C19- participants. Self-reported smell (**A,B**), taste (**C,D**), chemesthesis (**E,F**), and nasal obstruction (**G,H;** formulated as “How blocked was your nose?”) before and during respiratory illness in C19+ (darker shades) and C19-(lighter shades) participants. Ratings were given on 0-100 visual analog scales. Left panels (A,C,E,G) show mean values. Right panels (B,D,F,H) show distributions of the change scores (during minus before). Thicker sections indicate relatively more subjects (higher density of responses). The thick black horizontal bar indicates the median, the shaded area within each violin indicates the interquartile range. Each dot represents the rating of a single participant.

### Prediction of COVID-19 status from survey responses

Binary (yes/no) and categorical reported symptoms suggested that COVID-19 is more strongly associated with chemosensory than with non-chemosensory symptoms, including fever, cough, and shortness of breath, cardinal symptoms currently highlighted by the US Centers for Disease Control and Prevention (CDC)(**Figure 3A**). Using AUC to assess prediction quality (**Figure 3B**), we found that self-reported smell ability during illness, reported on a continuous scale, was the most predictive survey question for COVID-19 status(AUC=0.71). Changes in smell as a result of illness (the difference between smell ability during and before illness) was similarly predictive (AUC=0.69). Changes in taste ability (assessed via rating) were the next most predictive features (AUC=0.64-0.65) (**Figure 3B**). Models fit to the same data but with shuffled COVID-19 status consistently produced AUC∼0.5 for all features. The most predictive non-chemosensory symptom, sore throat (which was negatively associated with COVID-19) was substantially less predictive (AUC=0.58) than the top chemosensory symptoms. Nasal obstruction was not predictive (AUC=0.52). Responses given on a continuous scale were more predictive (AUC=0.71) than binary responses to parallel questions (e.g., **Appendix 1**, Question 13 versus 14, **FigureS5**) (AUC=0.60-0.62), likely because a continuous scale contains a greater amount of diagnostic information (**Figure S4**).

**Figure 3.**
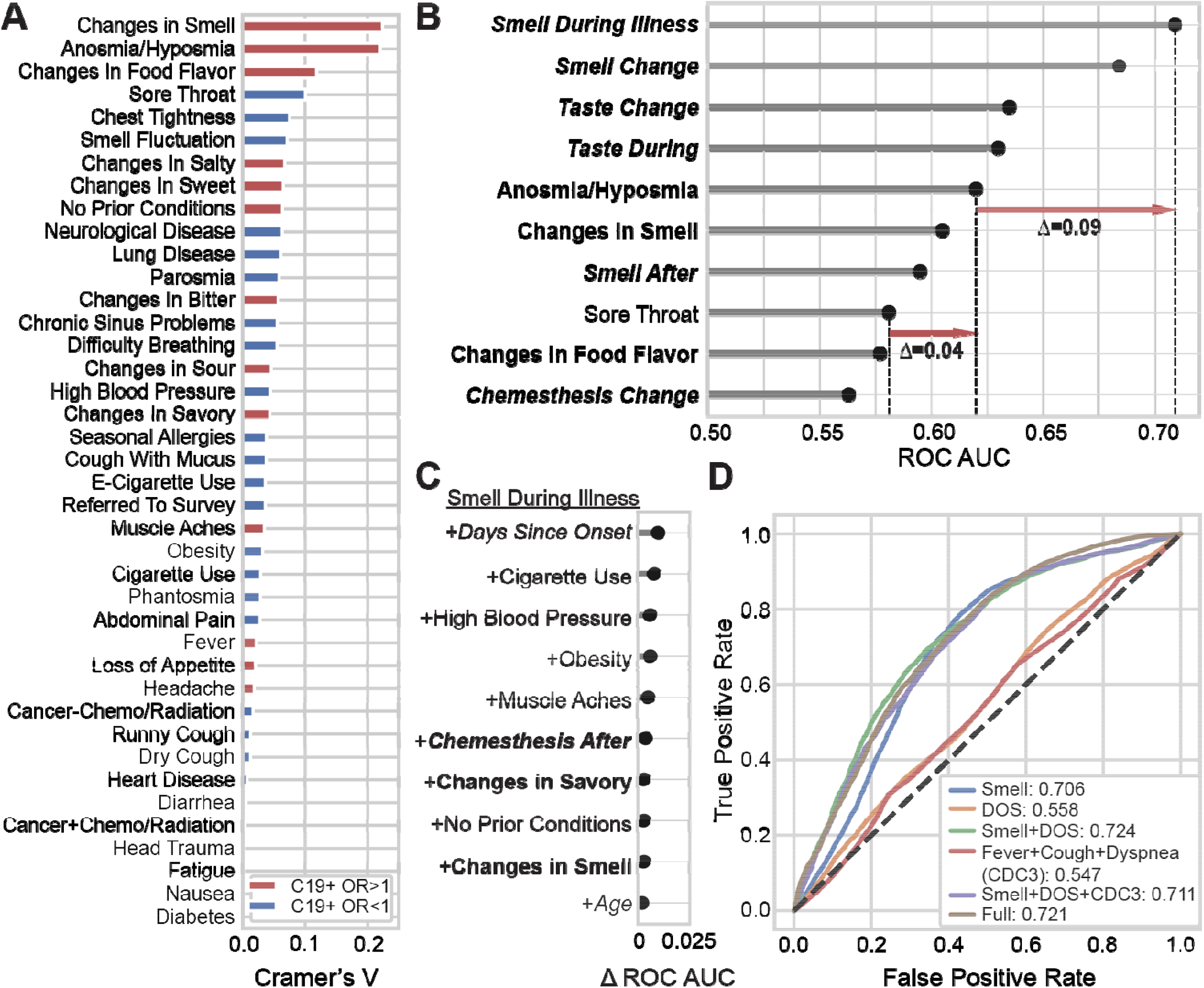
Smell loss is the strongest predictor of COVID-19 status. (**A**) A normalized measure of association (Cramer’s V) between binary or categorical responses on COVID-19 status. V=0 reflects no association between the response and COVID-19 status; V=1 reflects a perfect association; V>0.1 is considered a meaningful association. Features in red are positively associated with C19+ (odds ratio > 1); features in blue are negatively associated with C19+ (odds ratio < 1). (**B**) Logistic regression is used to predict COVID-19 status from individual features. Top-10 single features are ranked by performance (cross-validated <u>a</u>rea <u>u</u>nder the ROC <u>c</u>urve, AUC). Chemosensory-related features (bold) show greater predictive accuracy than non-chemosensory features (non-bold). Responses provided on the numeric scale (italic) were more informative than binary responses (non-italic). Red arrows indicate differences in prediction quality (in AUC) between features. (**C**) Adding features to “Smell During Illness” results in little improvement to the model; only **D**ays Since **O**nset of Respiratory **S**ymptoms (DOS) yields meaningful improvement. (**D**) ROC curves for several models. A model using “Smell during illness” (Smell Only, abbreviated “Smell” in figure) is compared against models containing this feature along with DOS, as well as models including the three cardinal CDC features (fever, dry cough, difficulty breathing). “Full” indicates a regularized model fit using 70 dozen survey features, which achieves prediction accuracy similar to the parsimonious model “Smell Only+DOS”.

Next, we examined which simple multi-feature model would best predict COVID-19 status. As some questions have highly correlated responses, the question most complementary to “Smell during illness” is unlikely to be one that carries redundant information. Adding “Days since onset of respiratory symptoms” (DOS) to “Smell during illness” (Smell Only) produced the largest incremental gain in predictive performance (AUC=0.72, +0.01 versus the Smell Only model) (**Figure 3C**).

We directly compared the Smell Only+DOS model to other candidate models. The Smell Only+DOS model (**Figure 3D**) yielded an equal or higher AUC than the model including the three cardinal CDC symptoms (AUC=0.55) or the full model using 70 features (AUC=0.72). Because the Smell Only+DOS model exhibits the same AUC as the full model it strikes a good balance between model parsimony and predictive accuracy for C19+. However, the Smell Only model also offers reasonable sensitivity of 0.85 (at specificity=0.51, cutoff=13 on the 100-point VAS) and/or specificity of 0.75 (at sensitivity=0.51, cutoff=1) as desired. By sharp contrast, fever has a sensitivity of only 0.54 with specificity of 0.49 and dry cough has sensitivity of 0.52 and specificity of 0.46.

### Recovery from smell loss

Recovery from smell loss was modest (approximately half the initial average loss) in C19+ participants with full or partial resolution of respiratory symptoms. Overall, self-reported, post-illness olfactory ability was still lower for C19+ (39.9±34.7) than C19- (52.2±35.2, p=2.8e-11, **Figure S6A**). However, the mean recovery of smell (after illness relative to during illness) was greater for C19+ (30.5±35.7) than C19-(24.6±31.9, p=0.0002, **Figure S6B**). A similar but smaller effect of COVID-19 status on recovery was observed for taste (**Figure S6C, D**), while little to no association with COVID-19 was observed for recovery of chemesthesis (**Figure S6E,F**) or nasal obstruction (**Figure S6G,H**). When illness-induced change in olfactory function (during minus before illness) and recovery of olfactory function (after minus during illness) were evaluated, we identified three respondent clusters: those self-reporting no loss of smell (Intact Smell), those reporting recovery from smell loss (Recovered Smell), and those reporting smell loss without recovery by up to 40 days (Persistent Smell Loss, **Figure 4, Table S3)**. Intact smell was reported by only 8.5% of the participants in the C19+ group but by 27.5% in the C19-group (p=3.8e-31). A greater proportion of C19+ participants were included in both the Recovered Smell group (C19+: 40.9%, C19-: 33.3%; p=4.9e-10) and the Persistent Smell Loss group (C19+: 50.7%, C19-: 39.2%; p=5e-5; **Figure 4A, B**). C19+ participants in both the Recovered Smell and Persistent Smell Loss clusters reported a similar extent of olfactory loss, irrespective of time since respiratory symptom onset. By contrast, the rate of self-reported smell recovery increased over time, with a plateau at 30 days (**Figure 4C**). Finally, DOS was the best predictor (AUC=0.62) between the Persistent Smell Loss and the Recovered Smell groups (**Figure S6A, Table S3**).

**Figure 4:**
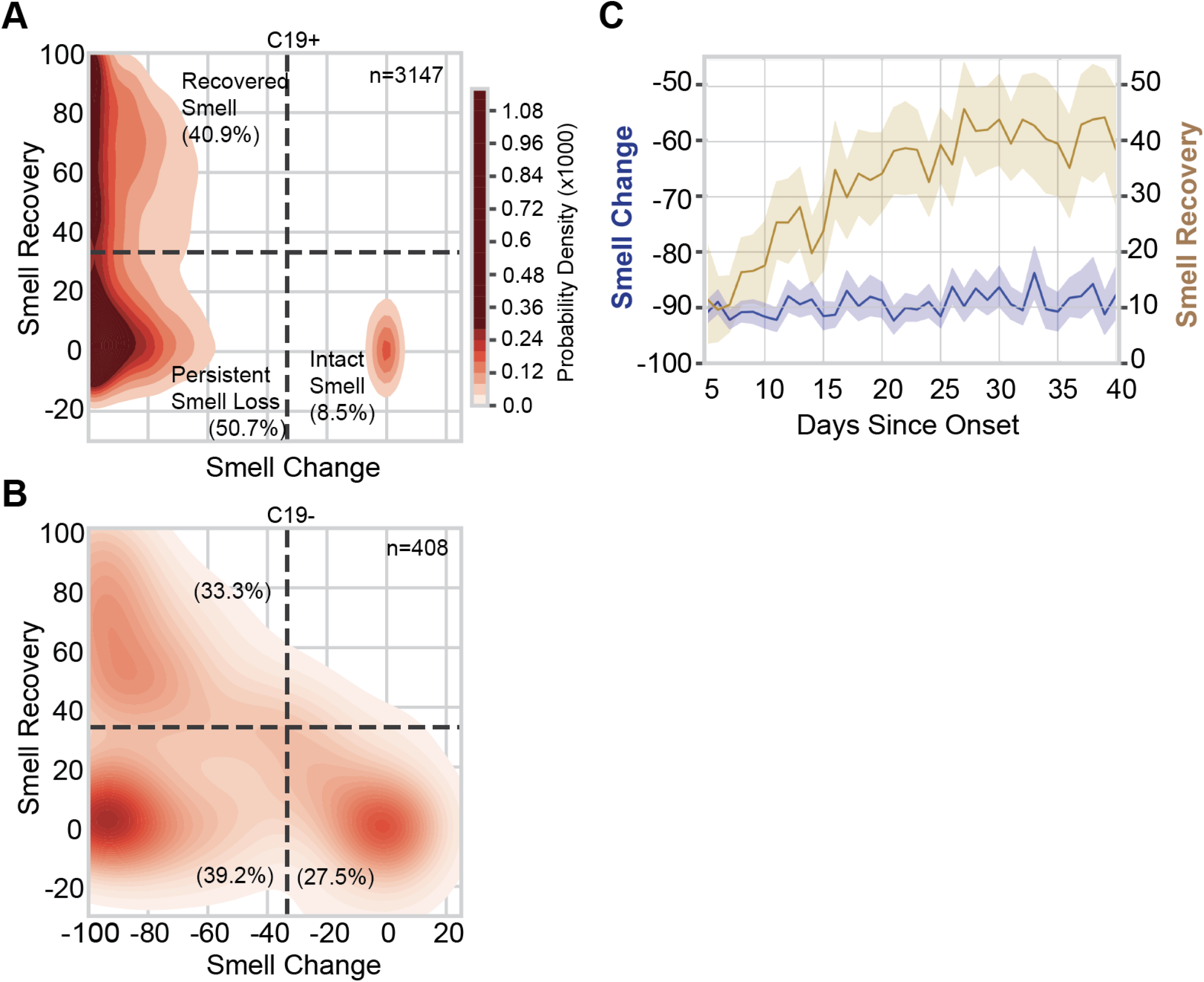
Smell loss, recovery, and time course. (**A, B)** Joint distribution of smell loss (during minus before illness ratings) and smell recovery (after minus during illness ratings) for C19+ (A) and C19- (B) participants. Darker color indicates a higher probability density; the color map is shared between (A) and (B); dashed lines are placed at a third of the way across the rating scale to aid visualization of the clusters. Severe smell loss that is either persistent (lower left) or recovered (upper left) was more common in C19+ than C19-. *n* indicates the number of participants in each panel. % indicates the percentage of participants of the given COVID status in each quadrant. (**C**) In C19+ participants who lost their sense of smell (Recovered Smell + Persistent Smell Loss), the degree of smell recovery (right y axis) increased over ∼30 days since onset of respiratory symptoms before plateauing; the degree of reported smell change (left y axis) did not vary in that window of observation. Solid lines indicate the mean of the measure, the shaded region indicates the 95% confidence interval.

### Simple screening for COVID-19: the **O**lfactory **D**eterminati**o**n **R**ating scale in COVID-**19** (ODoR-19) (136)

Our results indicate that a continuous rating of current olfactory function is the single best predictor of COVID-19 and improves the discrimination between C19+ and C19-over a binary question on smell loss. For example, the Smell Only model can reach a specificity of 0.83 at the low end of the VAS (sensitivity=0.36, cutoff=0). We propose here a numeric variation of the rating scale (0-10), the ODoR-19, that can be administered in person or via telemedicine to improve early COVID-19 screening for individuals without preexisting smell and/or taste disorders. Responses to the ODoR-19 scale ≤2 indicate high odds of COVID-19 positivity (4<OR<10, **Figure 5D**). An ODoR-19 response of 3 indicates a borderline risk (OR=1.2).

**Figure 5.**
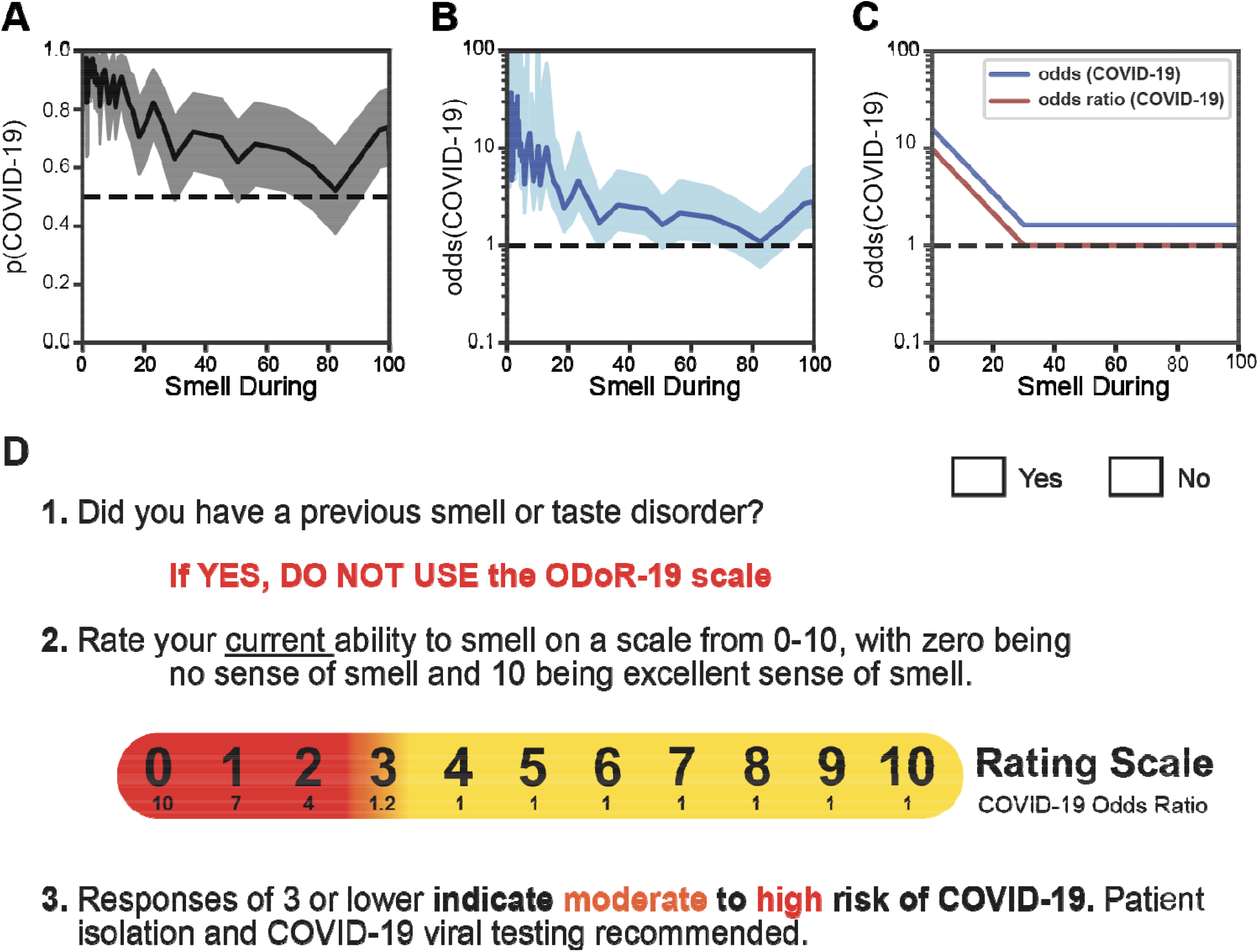
The odds of a COVID-19 diagnosis as a function of olfactory loss. (**A**) The solid line indicates the probability of a COVID-19 diagnosis as a function of “Smell during illness” ratings. The shaded region indicates the 95% confidence interval. (**B**) The solid line expresses the probability of a COVID-19+ diagnosis as a function of “Smell during illness” in odds (p/(1-p)); it is shown on a logarithmic scale. The shaded region indicates the 95% confidence interval. (**C**) Stylized depiction of change in the odds of a COVID-19 diagnosis and of the odds ratio. (**D**) The ODoR-19 screening tool. After healthcare providers or contact tracers have excluded previous smell and/or taste disorders such as those resulting from head trauma, chronic rhinosinusitis, or previous viral illness, the patient can be asked to rate their <u>current</u> ability to smell on a scale from 0-10, with 0 being no sense of smell and 10 being excellent sense of smell. If the patient reports a value below or equal to 3, there is a high (red) or moderate (orange) probability that the patient has COVID-19. Values in yellow (ratings above 3) cannot rule out COVID-19.

## Discussion

Self-reported smell loss was more common in C19+ than C19- participants, but present in both groups. The use of a VAS to assess olfactory loss better predicted COVID-19 status than using a binary question. We found that the best predictor of COVID-19-associated smell recovery, within the time frame captured by the survey (∼40 days), was days since onset of COVID-19.

The SARS-CoV-2 pandemic requires healthcare providers and contact tracers to quickly and reliably assess an individual’s COVID-19 risk, often remotely. Thus, reliable screening tools are critical to assess a person’s likelihood of having COVID-19 and to justify self-quarantine and/or testing recommendations. Indeed, some reports suggest that COVID-19-associated smell loss might be an indicator of disease severity.^2,20^ Current symptom criteria (e.g., fever, dry cough) are less specific than severe olfactory loss. Indeed, the value of our ODoR-19 tool lies in the high specificity of values ≤2 for indicating COVID-19 positivity, therefore representing a valuable addition to the current repertoire of COVID-19 screening tools. Those who receive a negative outcome from a COVID-19 viral test, yet report significant idiopathic smell loss, should be considered as high-priority candidates for COVID-19 re-testing.

Our online survey and sampling methodology likely selected participants with a heightened interest in smell and taste and/or their disturbances. This self-selection bias could be viewed as a limitation since the C19-group also showed chemosensory loss. However, finding difference between groups in a sample with a higher barrier for discriminating between C19+ and C19-supports the robustness of this tool when used in a typical clinical population; our collider bias analysis also suggests that our findings are likely conservative estimates (**Figure S1, Table S1**).

Our results suggest that chemosensory impairment has strong COVID-19 predictive value and is useful when access to viral testing is limited or absent. As with any self-report measure, veracity of self-reports cannot be guaranteed. However, the ability to screen individuals in real-time should outweigh this potential confound.^21^ While objective smell tests are the gold standard for assessing olfactory function,^22,23^ they are costly, time-consuming to administer, and can require in-person interactions with potentially infectious patients.^23,24^ By contrast, the ODoR-19 is free, quick, and can be administered in person or remotely. We cannot exclude that our C19-sample contains COVID-19 false negatives.^25^ However, self-reported smell during illness distinguishes between C19+ and C19-, but not between randomly shuffled cases, suggesting that the difference between C19+ and C19-, even in a sample with over-represented chemosensory dysfunction, is substantial and can be captured via self-report.

Approximately half of the participants in the C19+ group recovered their sense of smell within 40 days from the onset of respiratory symptoms. This suggests the presence of at least two subgroups of patients: one that recovers quickly (<40 days, 40.9%) and another that may present a more variable time course of recovery (50.7%). Since these data are collected before the full recovery of all symptoms, we cannot offer a complete picture of recovery from olfactory loss in COVID-19-positive individuals, but they align with other early reports.^26^ The COVID-19 pandemic will greatly increase the number of patients suffering from anosmia and other chemosensory disorders,^27^ conditions that significantly affect quality-of-life,^28,29^ dietary behavior,^30^ cardiovascular health,^31^ and mental health.^32,33^ Thus, it is necessary to prepare healthcare providers to address the long-term needs of these patients.

Based on our results, we propose the use of the ODoR-19 tool, a quick, free, and effective smell-based screening method for COVID-19. This 0-10 rating scale accurately predicts COVID-19 in individuals without pre-existing smell and taste disorders (e.g., from head trauma, chronic rhinosinusitis^34^). ODoR-19 combines the utility of a continuous scale with the ease and speed needed for a screening tool. ODoR-19 is safe for remote administration during an illness with high viral spread and can precede and complement viral testing. This tool will improve screening for patients with limited or no access to medical care around the globe.

## Data Availability

The data and annotated code will be available on GitHub (http://github.com/GCCR/GCCR002) upon publication.

http://github.com/rgerkin/GCCR002

## Acknowledgements

The authors wish to thank all study participants, patients, and patient advocates that have contributed to this project, including members of the AbScent Facebook group. The authors wish to thank Micaela Hayes, MD for her input on the clinical relevance of this project, Shannon Alshouse and Olivia Christman for their help in implementing the survey, Sara Lipson for her support, and the international online survey research firm YouGov for providing data gathered with the Imperial College London YouGov Covid 19 Behaviour Tracker.

## Supplementary Material

### Collider bias results in underestimation of an association between smell loss and COVID-19

As others have noted,^35^ collider bias, resulting from selection or conditioning on variables involved in the analysis, may result in the distorted association between COVID-19 and candidate symptoms or patient attributes. In the present sample, it is likely that we have selected for both a higher probability of COVID-19 and a higher probability of smell and taste disorders than the population at large. However, rather than leading to an overestimation of the positive correlation between smell loss and COVID-19, collider bias is expected to lead to an underestimation of this correlation (**Figure S1)**. If we consider the hypothetical scenario in which there is no association between smell loss and COVID-19 status in the general population, we would expect a distribution similar to that depicted in **Figure S1A**, where the correlation between the likelihood of smell change and likelihood of COVID-19 is r = 0. Based on our recruitment method, we expect that the participants who elected to complete the GCCR core questionnaire were likely to have COVID-19, smell loss, or both. We can simulate participant selection to reflect this hypothesis by censoring subjects which do not meet a fixed sum of smell loss and COVID-19 probabilities (i.e., the red dots are excluded from the calculation of the correlation; **Figure S1B**). As a result, the estimated correlation between smell loss and COVID-19 status originating from a population with r = 0 would be negative (**Figure S1B)**. A similar scenario would manifest if the association between smell loss and COVID-19 status in the general population is positive (**Figure S1C**). Again, simulating the removal of participants with low likelihood of having COVID-19 and/or reporting smell loss would result in a bias of the estimated correlation towards more negative values (**Figure S1D**). This collider bias indicates that the positive correlation between smell loss and C19+ is underestimated in the present sample. Indeed, a direct comparison of the binary (y/n) smell loss questions in the two empirical samples yields an C19 odds ratio of 5.96 in the YouGov sample (**Table S1**) but only 4.89 for GCCR. Therefore, our analyses represent a conservative scenario for the prediction of C19+ and C19-based on chemosensory alterations.

**Figure S1.**
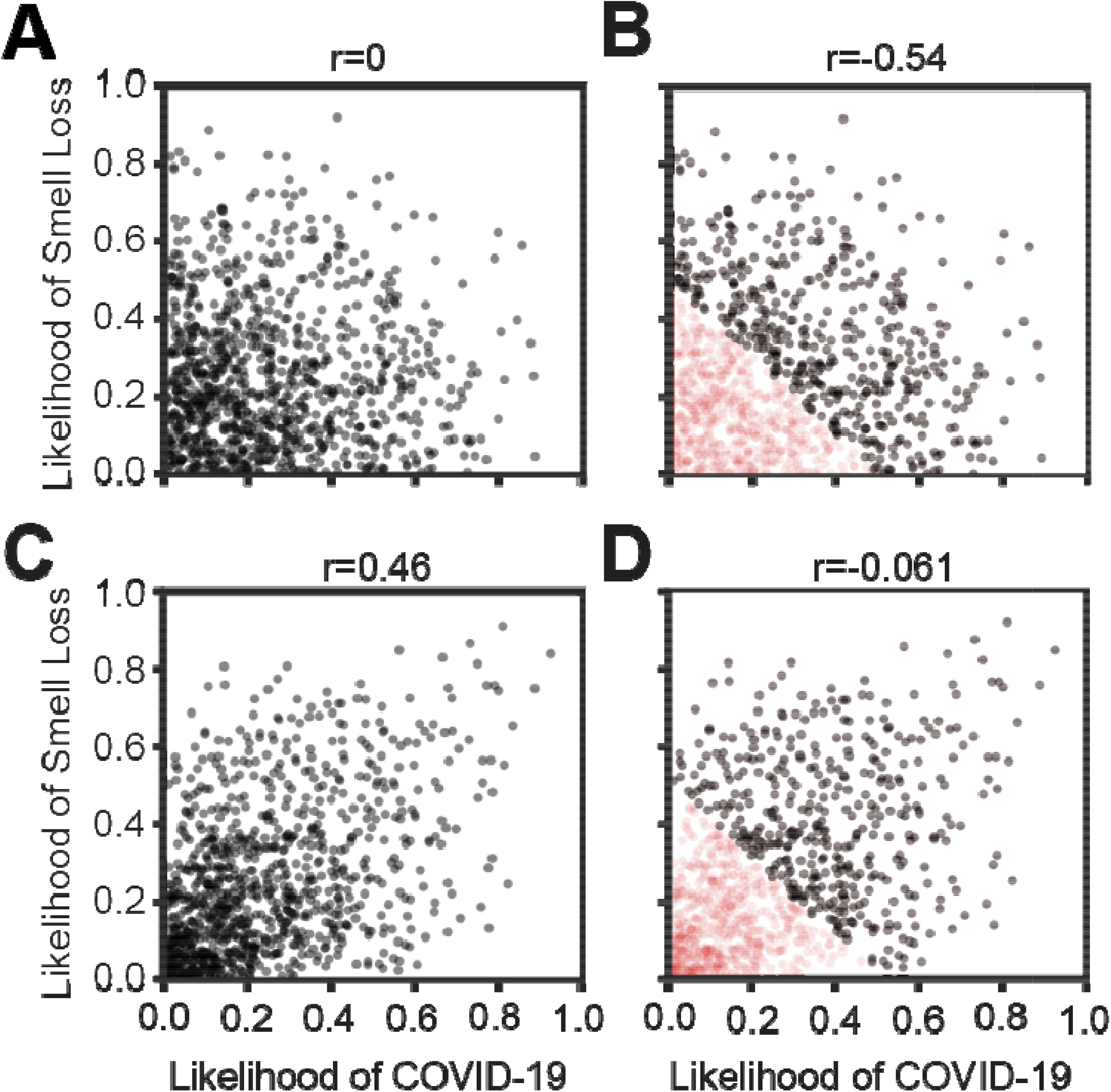
Collider bias leads to underestimation of the positive correlation between smell loss and COVID-19 positive ststus. **A)** Hypothetical scenario depicting no relationship between smell change and likelihood of COVID-19 positive status. Black dots indicate individual potential subjects, each of whom has a latent likelihood of COVID-19 and of smell loss. **(B)** Hypothetical scenario depicting the emergence of a negative correlation between smell change and likelihood of COVID-19 positive status following a baseline lack of correlation, if participants with greater smell loss and/or COVID-19 positive are preferentially included in the sample. Red dots indicate subjects not observed due to this selection bias; subjects observed remain in black. **(C)** Hypothetical scenario depicting a positive relationship between smell change and likelihood of COVID-19 positive status. **(D)** Hypothetical scenario depicting the emergence of a negative correlation between smell change and likelihood of COVID-19 positive status following a positive baseline correlation, if participants with greater smell loss and/or COVID-19 positive are preferentially included in the sample.

**Table S1.** Comparison with the representative YouGov database shows that the GCCR sample underestimates the positive correlation between smell loss and COVID-19 positive status.

|  | N | % C19+ | % C19- | OR<br>(C19+ vs C19-) | OR<br>(C19+ vs Not Tested) | OR<br>(C19- vs Not Tested) | p(Smell Loss \| C19+) |
| --- | --- | --- | --- | --- | --- | --- | --- |
| **Global** | 109395 | 0.51 | 3 | 5.96 | 49.7 | 8.33 | 0.33 |
| **Brazil** | 4167 | 0.91 | 5.4 | 8.39 | 33.9 | 4.04 | 0.47 |
| **Canada** | 5524 | 0.47 | 3.4 | 7.76 | 71.6 | 9.22 | 0.35 |
| **Denmark** | 5839 | 0.36 | 5.7 | 9.35 | 77.8 | 8.32 | 0.33 |
| **Finland** | 5927 | 0.32 | 1.9 | 1.85 | 27.8 | 15.1 | 0.21 |
| **France** | 9820 | 0.46 | 1.7 | 7 | 57.9 | 8.27 | 0.29 |
| **Germany** | 9468 | 0.49 | 2.1 | 4.56 | 94.2 | 20.7 | 0.37 |
| **Italy** | 9790 | 0.33 | 2.8 | 9.17 | 50.8 | 5.54 | 0.22 |
| **Mexico** | 5840 | 0.24 | 3.5 | 8.91 | 27.6 | 3.1 | 0.21 |
| **Netherlands** | 3822 | 1.2 | 2.7 | 2.73 | 16.1 | 5.92 | 0.3 |
| **Norway** | 5794 | 0.86 | 4.4 | 5.82 | 51.2 | 8.8 | 0.32 |
| **Spain** | 9789 | 0.37 | 3.5 | 5.81 | 43.5 | 7.48 | 0.33 |
| **Sweden** | 9741 | 0.44 | 2 | 3.69 | 38 | 10.3 | 0.35 |
| **United Kingdom** | 13565 | 0.15 | 1.2 | 23.1 | 108 | 4.66 | 0.5 |
| **United States** | 10309 | 1.2 | 4.8 | 4.61 | 54.7 | 11.9 | 0.34 |

### How representative is the GCCR sample?

As with most COVID-19 studies,^19^ the sample studied here is not representative of the general population. To better understand the extent to which this is the case, we computed a cross-correlation between GCCR and YouGov data.^36^ These data were aligned by weighting YouGov samples to achieve an identical survey date distribution to the GCCR samples. Specifically, GCCR survey dates were converted to a YouGov “week number” because YouGov surveys only weekly. The distribution of week numbers was computed for each country in the GCCR data. The YouGov data for the same country was then weighted by week number to match the corresponding GCCR distribution for that country. So, for example, if a country had 10 GCCR survey responses in week 1 of the YouGov survey period, and 30 in week 2 of that period, then the YouGov data in week 1 would be weighted at 25% and in the YouGov data in week 2 at 75%. This procedure was applied independently for each country, and the weights were used to compute a weighted mean COVID-19-positive rate for each country from the YouGov data. This was then directly compared against the raw COVID-19-positive rate for each country in the GCCR data. A lag (x-axis value in **Figure S2**) of 0 exactly reflects the above description. Other values of the lag indicate that the alignment was shifted: for example, a lag of one week means that the hypothetical GCCR responses above would be weighted 25/75 towards weeks 2 and 3, instead of weeks 1 and 2. Under the hypothesis that the COVID-19-positive rates in the two surveys are related, but may have different temporal dynamics, changing the lag allows these dynamics to be estimated. **Figure S2** depicts the country-wise correlation in participants with a positive COVID-19 test results (C19+) fraction between the two datasets, as a function of the lag between GCCR survey date and YouGov survey date. The country-wise C19+ fraction is correlated (r ∼ 0.45) when responses from the same calendar week are aligned, but diminishes outside of that window, showing both surveys capture a similar within-country temporal component of the epidemic.

**Figure S2.**
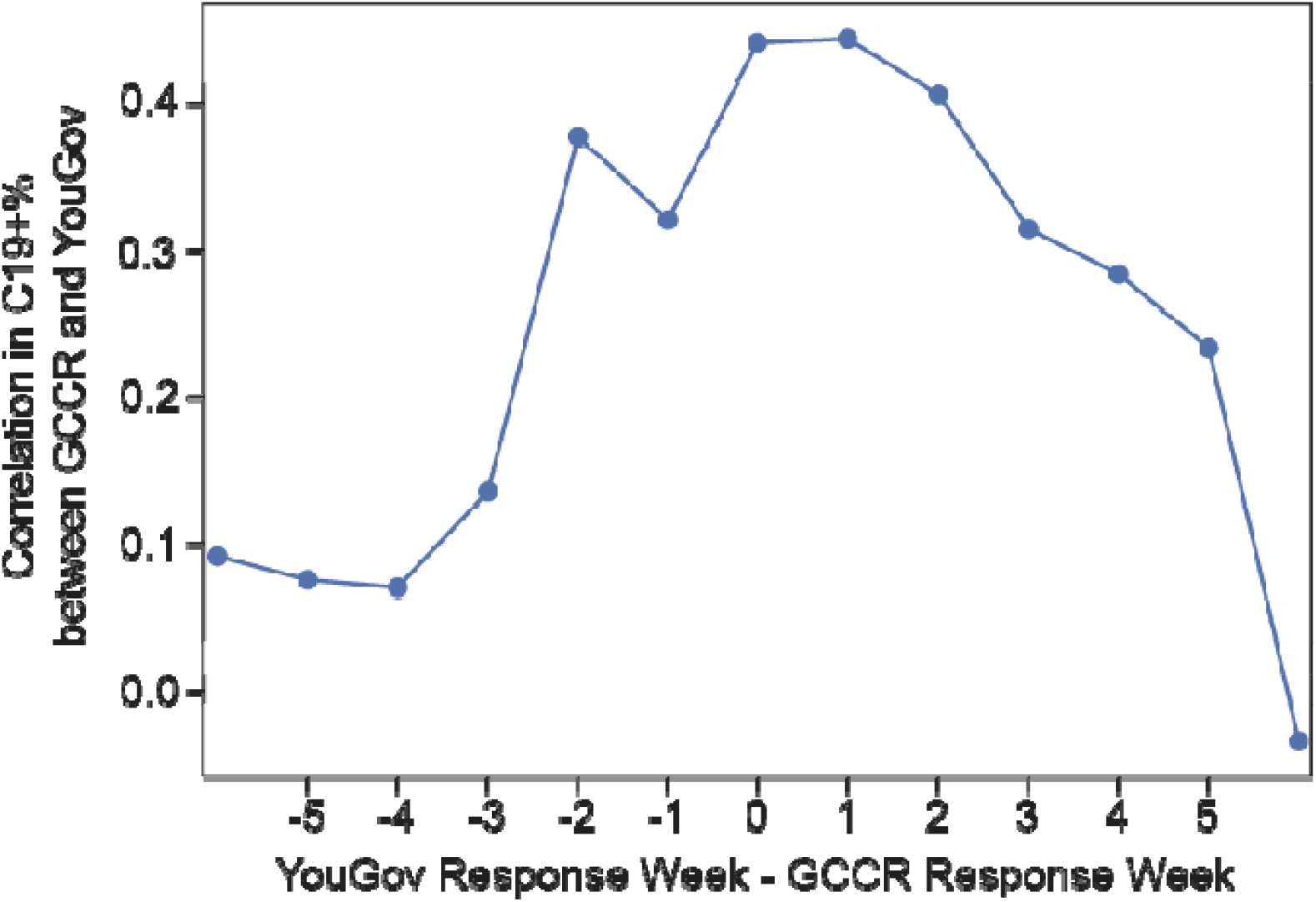
COVID-19 status in the GCCR cohort is correlated with a representative YouGov sample.

### Sample description

Based on responses to question 7 of the GCCR survey (“Have you been diagnosed with COVID-19?”, **Appendix 1**), participants can be split into six groups (see **Figure 1**). Participants who responded with Option 2 (“Yes – diagnosed with viral swab”) or 3 (“Yes – diagnosed with another lab test”) were classified as C19+; participants who responded with option 5 (“No – I had a negative test, but I have symptoms”) were classified as symptomatic C19-; participants who responded with option 4 (“No – I was not diagnosed, but I have symptoms”) were classified as C19 Unknown; participants who responded with option 6 (“No – I do not have any symptoms”), with option 7 (“Don’t know”), or with option 8 (“Other”) were classified as undefinable and excluded from the final analyses. To replicate our previous findings,^8^ we first compared individuals newly included in the GCCR dataset (responses from 14 May to 2 July, 2020, replication sample in **Figure 1**) with COVID-19 who were lab tested and those who were diagnosed by a clinician based on the self-reported quantitative changes in smell, taste, chemesthesis, and nasal obstruction (**Figure S3)**. Participants with lab-test confirmed C19+ did show slightly greater chemosensory deficits than did those diagnosed with C19+ clinically, but the difference was not clinically meaningful (smell: 4.4±28.6, p=2.7e-13) (**Figure S3, Table S2**). We then focused our descriptive and predictive analyses of participants who received a positive (C19+) or a negative (C19-) lab test for COVID-19. We also computed descriptive and predictive analysis for the C19+ subsample who reported partial or full signs of recovery from their recent respiratory illness. Lastly, the unknown group was originally hypothesized as similar to the C19-group. Yet the ratings of smell ability during illness suggest that the majority of these participants has a smell profile closer to C19+ than C19- (**Figure S4**). To maximize the validity of the COVID-19 diagnosis in our sample, we therefore excluded the C19 Unknown group from further analyses.

**Figure S3.**
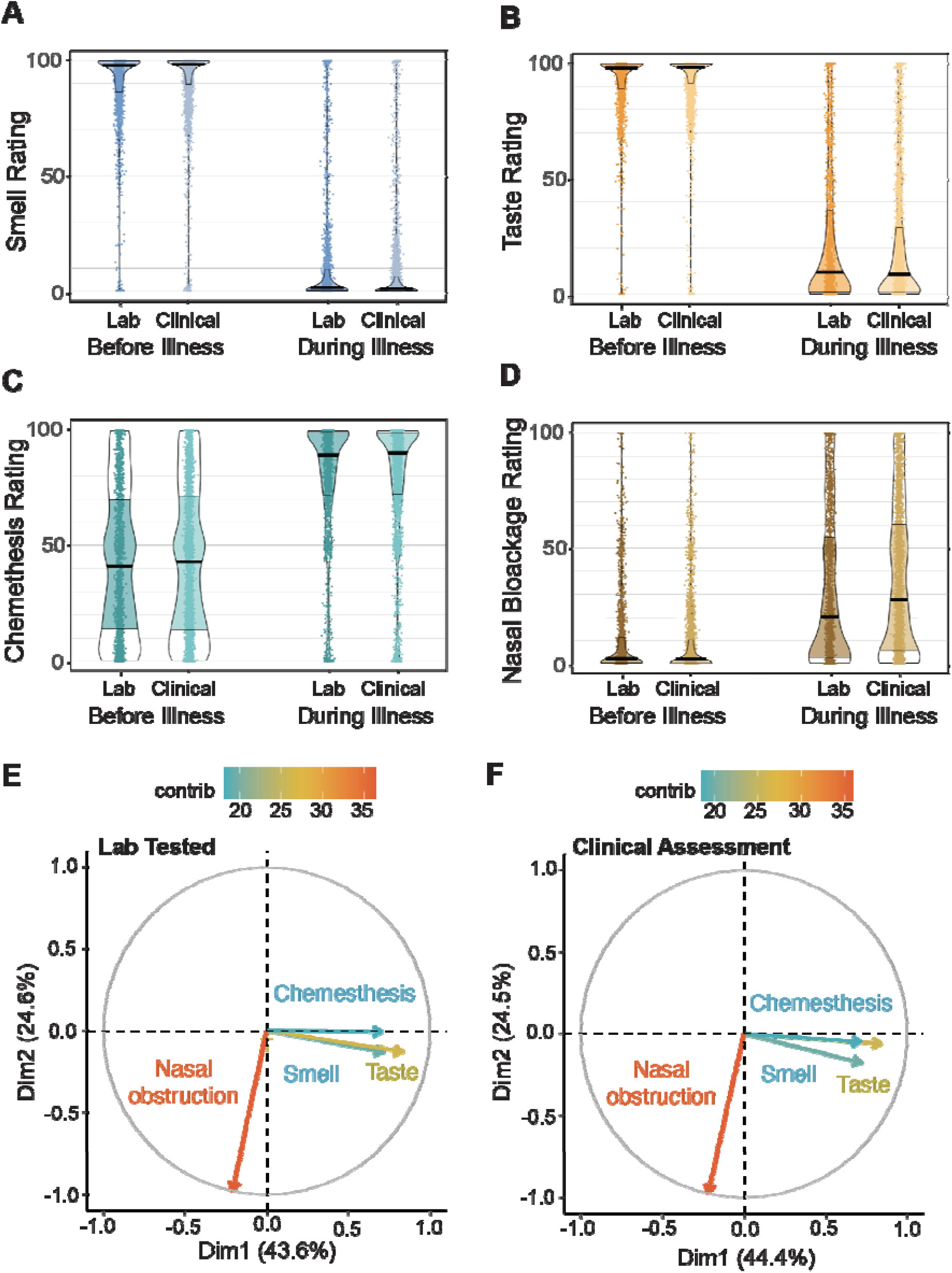
This figure describes a pre-registered replication of Parma et al, 2020 and includes only new data collected between May 14th and July 3rd 2020 via the GCCR survey. (**A-D**) Changes in smell (A), taste (B), chemesthesis (C) and nasal blockage (D) during versus before in COVID-19-positive individuals (Groups 1, 2 and 3, see Figure 1). All subjects had a COVID-19-positive status either via lab test (darker shades) or via clinical assessment (lighter shades). (**E-F**) Principal component analysis shows that smell, taste, and chemesthesis changes in both the lab test (E) and clinical assessment (F) groups) were orthogonal to blocked nose changes, i.e., the three chemosensory changes were highly correlated across subjects whereas blocked nose changes were mostly uncorrelated.

**Figure S4.**
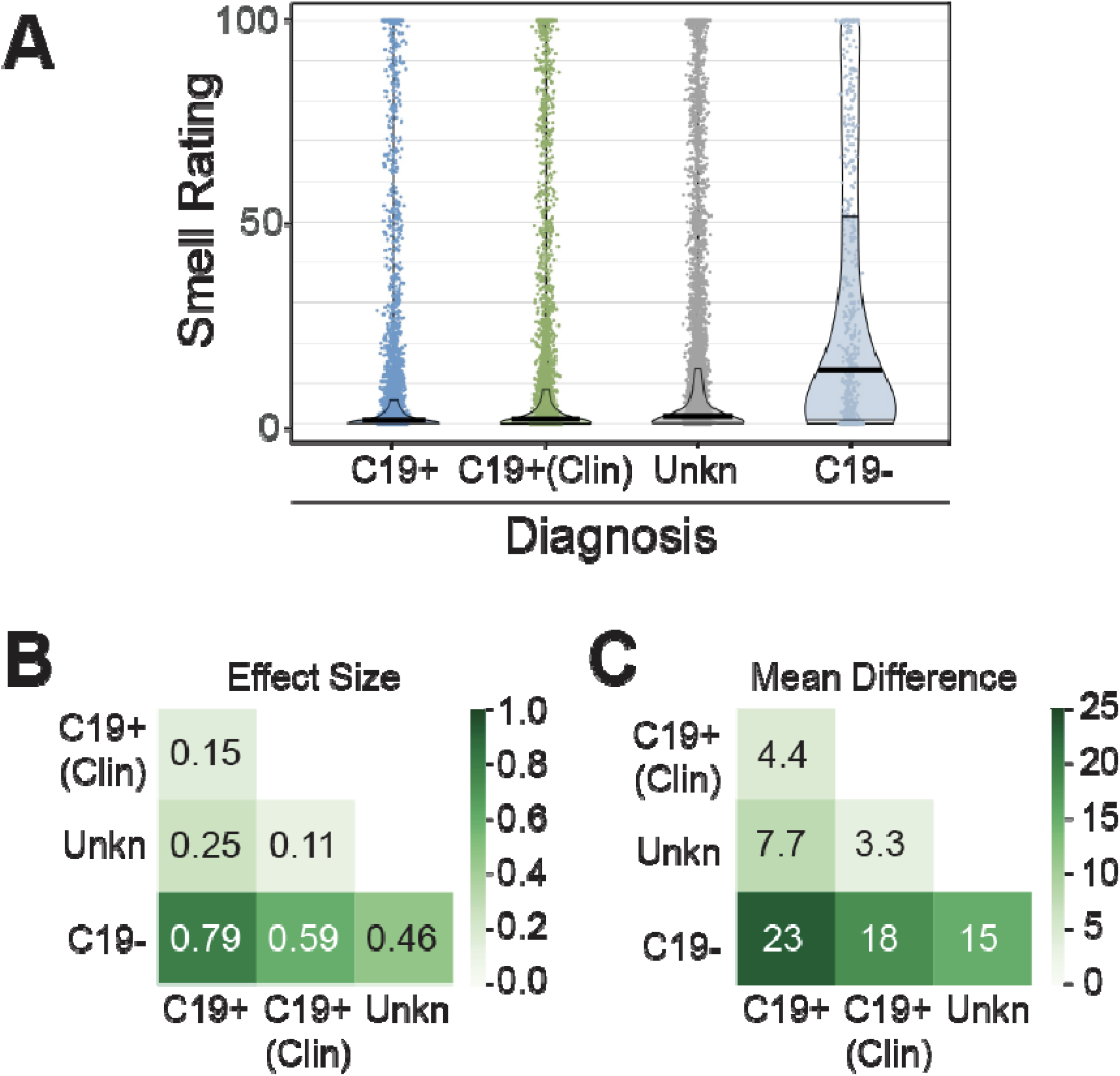
(**A**) Self-reported smell change and comparison of smell change between four diagnosis groups: Positive COVID-19 lab-test (C19+), positive COVID-19 clinical assessment (C19+ (Clin)), COVID-19 Unknown (Unkn; lack of clinical and lab test diagnosis, but reported symptoms), and negative COVID-19 lab test (C19-). Solid horizontal lines reflect the median; dashed lines reflect the quartiles. (**B, C**) Differences between groups, in terms of (B) effect size (Cohen’s D) and (C) means (on a 0-100 scale).

### Replication of previous analyses

The replication of Parma et al.^8^ used the same Bayesian linear regression approach with Cauchy prior [r = sqrt(2)/2]. This approach is appropriate for estimating the strength of the evidence in support of the alternative hypothesis: the clinical assessment and the lab test C19+ groups show similar smell, taste, chemesthesis and nasal obstruction changes before vs. during the illness. The interpretation of the Bayes factors BF follows the classification scheme proposed by Lee and Wagenmakers^37^ and adjusted from Jeffreys^38^, which considers BF > 3 as moderate evidence, BF > 10 as strong evidence, BF > 30 as very strong evidence and BF > 100 as extreme evidence for H_0_ or H_1_.

**Table S2.** Differences between lab-tested and clinically-assessed COVID-19-positive participants on changes in smell, taste, chemesthesis and nasal blockage.

|  | Smell Change | Taste Change | Chemesthesis Change | Change in Nasal Blockage |
| --- | --- | --- | --- | --- |
| $\Delta$ | -4.4 | -3.4 | 0.37 | 3.9 |
| $\sigma$ | 0 | 0 | 37 | 33 |
| seΔ | -0.048 | -0.037 | 0.0041 | 0.043 |
| D | -0.15 | -0.1 | 0.01 | 0.12 |
| p | 2.70E-13 | 2.00E-06 | 0.38 | 4.00E-09 |

Change means the rating "before" illness minus the rating "during" illness on the 0-100 visual-analog scale. $\Delta$ indicates the mean difference in change between lab-test and clinically-assessed COVID-19-positive subjects, while $\sigma$ indicates the standard deviation. $D$ indicates effect size (Cohen's D). $p$ indicates p-value from a Mann-Whitney U-test. In contrast to the prediction of the pre-registration, we found statistically significant differences between groups. However, the effect sizes are small and thus unlikely to be of practical importance.

### Chemosensory characterization of C19+ and C19-

We asked how accurately COVID-19 status could be predicted from the survey responses. The data matrix had strictly non-negative values and was normalized (column-wise min=0, max=1) to apply regularization in an equitable fashion across features and give regression coefficients the same interpretation for each feature. Compared with the main text, models with similar AUC values (but with non-zero coefficients for additional, likely spurious features) were obtained for smaller values of α, and inferior results for larger ones (which contained fewer or no non-zero coefficients). Quantitatively similar AUC values were obtained for other models predicting COVID-19 status using multiple features including ridge regression and random forest, but L1-regularized logistic regression consistently produced sparser models with comparable cross-validation accuracy. Each logistic regression model included an intercept term and one or more normalized features. Each model attempted to predict, using the value of the response to a single question (and an additive constant), whether a subject reported a C19+ or C19-status. Coefficients in a logistic regression model can be interpreted as changes in odds, or as odds ratios when two values are compared. Each ROC curve -- constructed using predictions on holdout test sets and concatenated over these test sets -- summarizes the tradeoff between sensitivity (fraction of C19+ cases correctly identified) and specificity (fraction of C19-cases correctly identified) as the threshold value for the predictor is varied.

### Value of using a scale rather than a binary response to detect C19+

We quantified the information entropy for each survey question used the following standard equation: 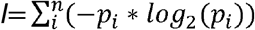 evaluated over the *n* response options. Re-binning to mimic new scales was achieved by dividing response values by a constant and rounding to the nearest integer. Relative mutual information was calculated by computing the mutual information between survey response and COVID status based on the following standard equation: 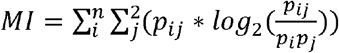 where survey response options are indexed with *i* and the C19+/C19-status (two possible values) are indexed with *j*, and then dividing by the entropy available from that same C19 status distribution, calculated using the first equation. Results indicate that soliciting responses on either a continuous 100-point scale or a downsampled 10-point numeric version of the scale is more informative about symptoms themselves and about COVID-19 status (given the symptoms) than soliciting binary responses (**Figure S5**).

**Figure S5.**
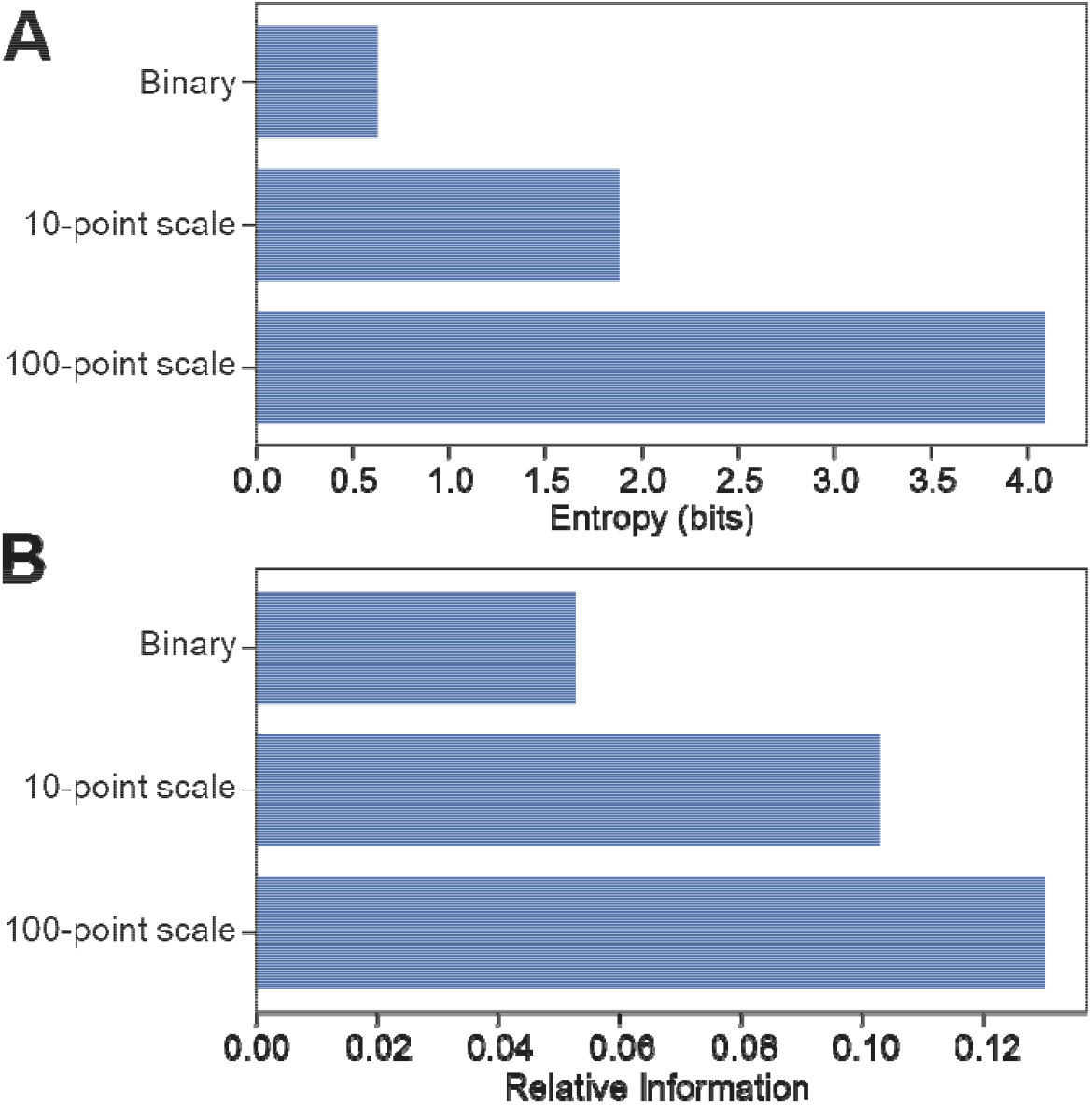
**(A)** Relative information available from the distribution of responses to the two primary “Smell” survey questions. *Binary* refers to the yes/no question about symptomatic smell loss. A relative information of 1 would correspond to a question whose response is perfectly informative about COVID-19 status. By contrast, a similar question asked on a numeric scale (0-100, the original scale; or a hypothetical 10-point scale obtained by rounding responses) contains substantially more information due to the resolution of the scale. A 10-point scale may be familiar from clinical self-reports of pain. (**B)** The relative mutual information about COVID status contained in the survey response is also higher for the full numeric scale or the hypothetical 10-point scale than for the binary question.

### Prediction of recovery from COVID-19-associated smell loss

We applied the same predictive modeling framework used in **Figure 4** to try to predict smell recovery in C19+ participants. In other words, we asked which survey responses predicted that a subject would fall into the Recovered Smell rather than the Persistent Smell Loss cluster, given both smell loss during the disease and C19+ status. The only predictive feature of any practical significance was “Days Since Onset” of respiratory symptoms (AUC=0.62), indicating that those who experienced their first respiratory symptoms less recently are more likely to have Recovered Smell (**Figure S6A**). Adding additional features to the model provided modest improvement (AUC=0.65 for the optimal model), but overall it was difficult to predict whether a C19+ participant would exhibit Recovered Smell or Persistent Smell Loss based on the data available (**Figure S6B**). **Table S3** includes the means and SD by recovery group for C19+ and C19-participants.

**Figure S6.**
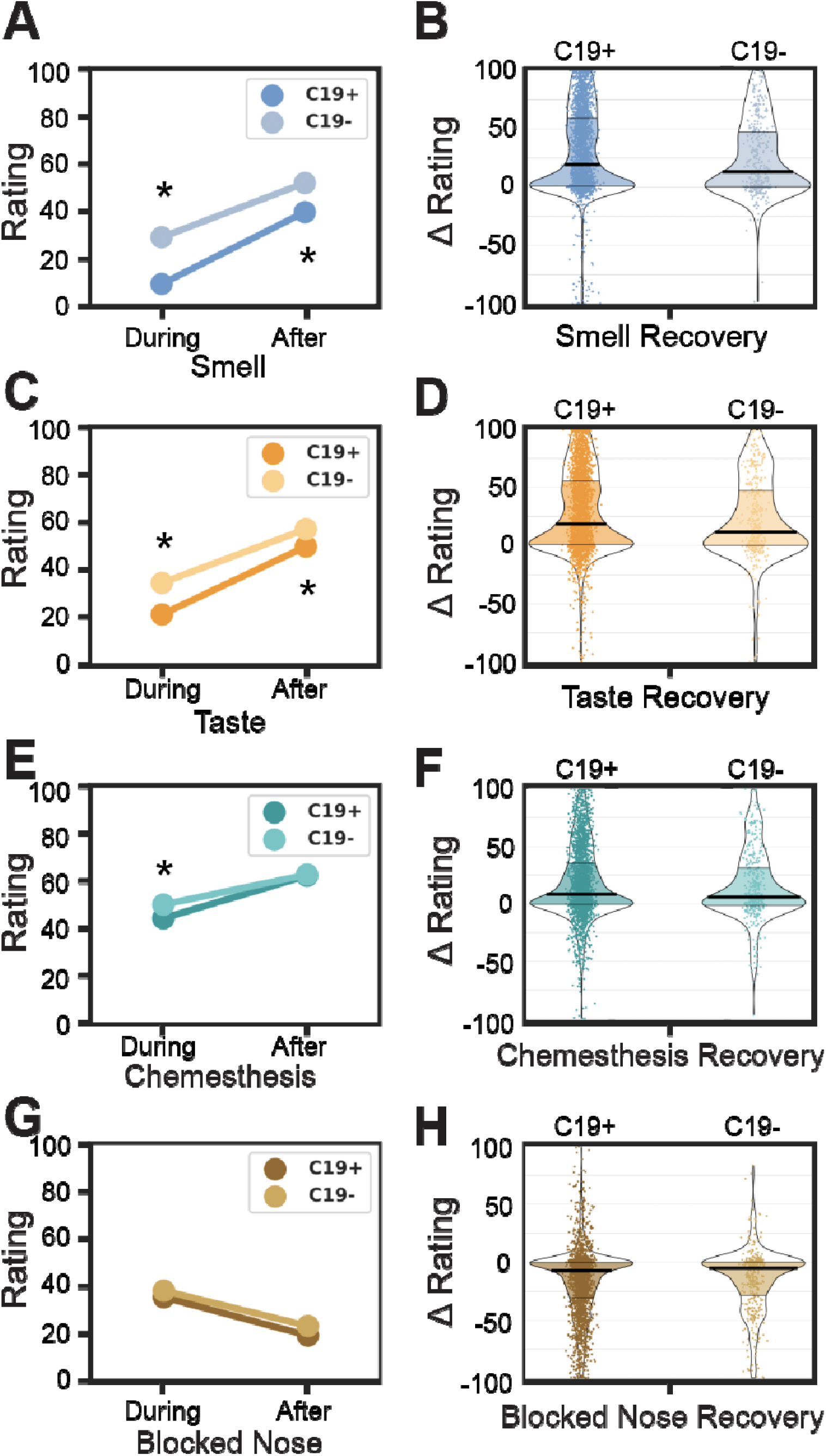
COVID-19 recovery. Similar to Figure 1, but self-reported smell (**A,B**), taste (**C,D**), chemesthesis (**E,F**), and nasal blockage(**G,H**) during and after respiratory illness in C19+ (darker) versus C19- (lighter). (A,C,E,G) mean values during and after respiratory illness, respectively. (B,D,F,H) Change (after minus during) as a distribution over subjects.

**Table S3.** Main chemosensory and relevant demographic features in the three clusters of recovering C19+ and C19- participants.

|  | Intact Smell |  |  | Persistent Smell Loss |  |  | Recovered Smell |  |  |
| --- | --- | --- | --- | --- | --- | --- | --- | --- | --- |
|  | C19+ | C19- | p | C19+ | C19- | p | C19+ | C19- | p |
| **Taste Change** | -34 ± 39 | -21 ± 30 | 0.011 | -75 ± 28 | -68 ± 32 | 0.0018 | -75 ± 29 | -69 ± 31 | 0.0043 |
| **Chemesthesis Change** | -17 ± 32 | -5.9 ± 25 | 0.016 | -40 ± 37 | -42 ± 38 | 0.2 | -36 ± 37 | -37 ± 39 | 0.41 |
| **Changes In Sweet** | 0.31 ± 0.46 | 0.19 ± 0.39 | 0.0067 | 0.46 ± 0.5 | 0.4 ± 0.49 | 0.067 | 0.47 ± 0.5 | 0.45 ± 0.5 | 0.36 |
| **Changes In Sour** | 0.27 ± 0.45 | 0.14 ± 0.35 | 0.0037 | 0.43 ± 0.5 | 0.42 ± 0.5 | 0.42 | 0.4 ± 0.49 | 0.42 ± 0.5 | 0.29 |
| **Smell Change** | -3.6 ± 14 | -8.1 ± 12 | 0.022 | -89 ± 13 | -82 ± 19 | 1.50E-07 | -89 ± 13 | -83 ± 17 | 4.20E-05 |
| **Phantosmia** | 0.09 ± 0.29 | 0.11 ± 0.31 | 0.35 | 0.09 ± 0.28 | 0.11 ± 0.31 | 0.21 | 0.08 ± 0.28 | 0.10 ± 0.3 | 0.32 |
| **Days Since Onset** | 28 ± 18 | 27 ± 17 | 0.35 | 27 ± 20 | 33 ± 26 | 0.0039 | 32 ± 17 | 33 ± 20 | 0.32 |
| **Parosmia** | 0.15 ± 0.36 | 0.15 ± 0.36 | 0.48 | 0.09 ± 0.28 | 0.12 ± 0.33 | 0.063 | 0.12 ± 0.33 | 0.15 ± 0.36 | 0.19 |
| **Changes In Savory** | 0.18 ± 0.39 | 0.16 ± 0.37 | 0.32 | 0.4 ± 0.49 | 0.36 ± 0.48 | 0.17 | 0.32 ± 0.47 | 0.35 ± 0.48 | 0.26 |
| **Onset Day** | 95 ± 24 | 91 ± 23 | 0.078 | 1e+02 ± 29 | 95 ± 30 | 0.00036 | 90 ± 21 | 91 ± 27 | 0.47 |
| **Taste Recovery** | 15 ± 33 | 13 ± 28 | 0.43 | 9.2 ± 23 | 6 ± 29 | 0.07 | 55 ± 30 | 50 ± 31 | 0.043 |
| **Changes In Bitter** | 0.27 ± 0.45 | 0.17 ± 0.38 | 0.018 | 0.43 ± 0.5 | 0.4 ± 0.49 | 0.2 | 0.41 ± 0.49 | 0.4 ± 0.49 | 0.41 |
| **Changes In Salty** | 0.36 ± 0.48 | 0.21 ± 0.41 | 0.0026 | 0.47 ± 0.5 | 0.42 ± 0.49 | 0.12 | 0.47 ± 0.5 | 0.44 ± 0.5 | 0.23 |
| **Fraction Women** | 0.63 ± 0.48 | 0.79 ± 0.41 | 0.0017 | 0.76 ± 0.43 | 0.77 ± 0.42 | 0.38 | 0.73 ± 0.44 | 0.75 ± 0.43 | 0.32 |
| **Anosmia/Hyposmia** | 0.43 ± 0.5 | 0.14 ± 0.35 | 3.50E-08 | 0.94 ± 0.23 | 0.86 ± 0.35 | 1.40E-05 | 0.88 ± 0.32 | 0.86 ± 0.35 | 0.23 |
| **Chemesthesis Recovery** | 8.3 ± 27 | 8.3 ± 24 | 0.24 | 9.5 ± 26 | 10 ± 27 | 0.41 | 29 ± 33 | 25 ± 38 | 0.25 |
| **Smell Recovery** | -3.8 ± 39 | 4.4 ± 22 | 0.14 | 6.3 ± 10 | 6.3 ± 11 | 0.22 | 67 ± 19 | 62 ± 18 | 0.007 |
| **Age** | 44 ± 13 | 44 ± 13 | 0.33 | 41 ± 12 | 42 ± 12 | 0.092 | 40 ± 12 | 43 ± 12 | 0.0015 |

## APPENDIX 1

### GCCR core questionnaire

The core questionnaire of the Global Consortium for Chemosensory Research (GCCR) has been deployed in Compusense Cloud in 32 languages. The questionnaire was published previously^8^ and also appears in the NIH Office of Behavioral and Social Sciences Research (OBSSR) research tools for COVID-19.^39^ Responses to the GCCR core questionnaire in 23 languages were collected between April 7 and July 2, 2020 and included in the final dataset, on which we conducted the analyses reported in this paper.

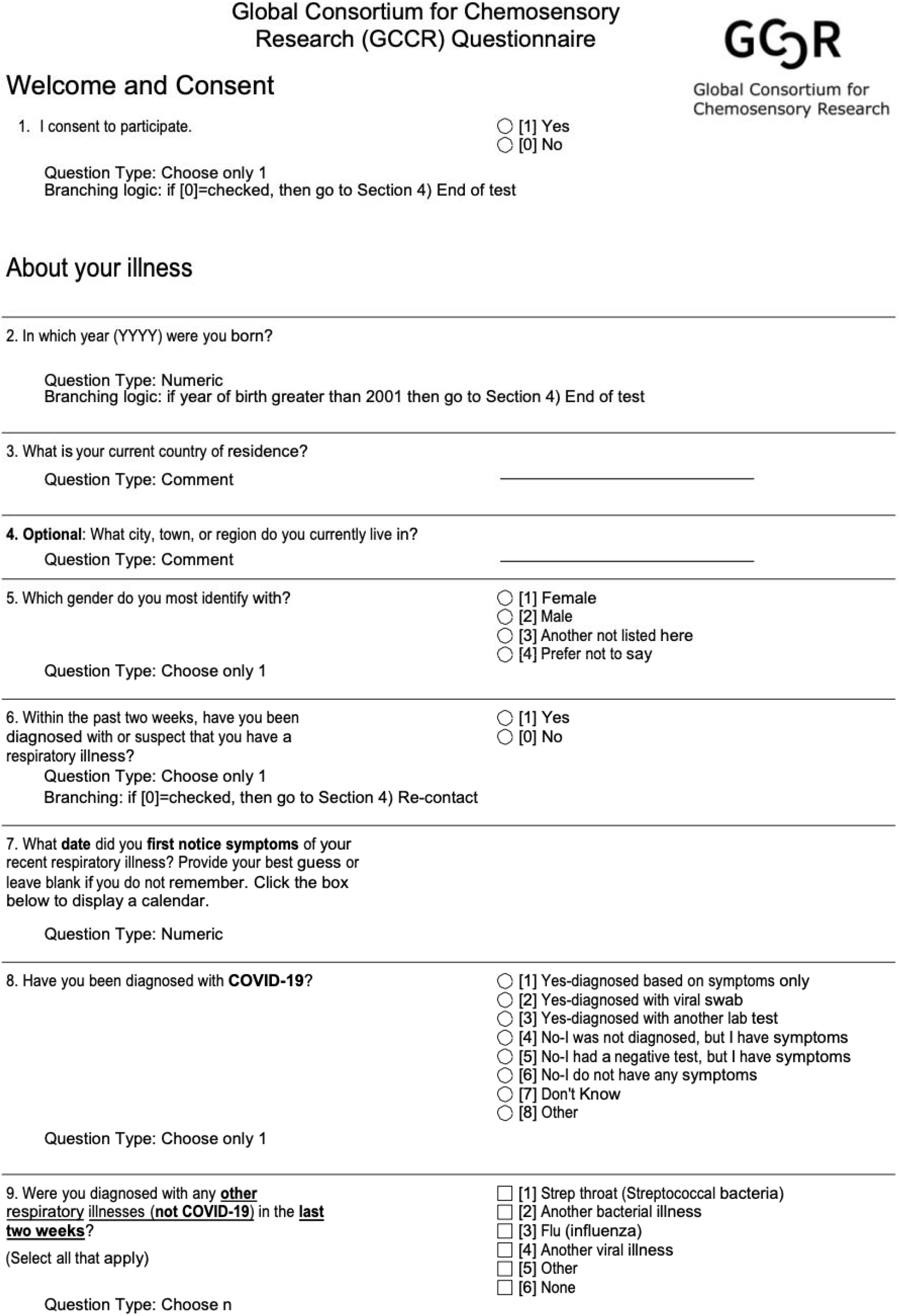

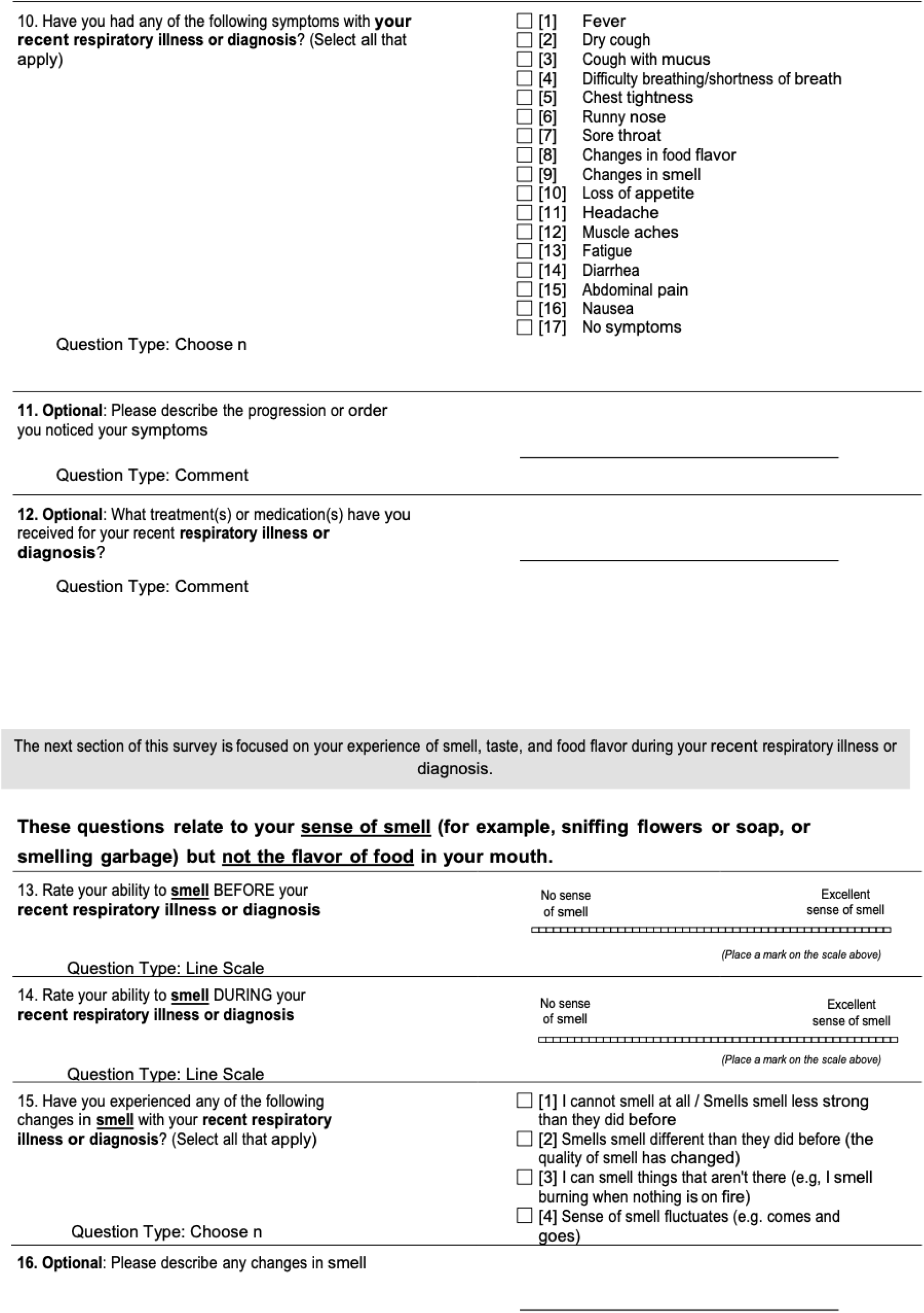

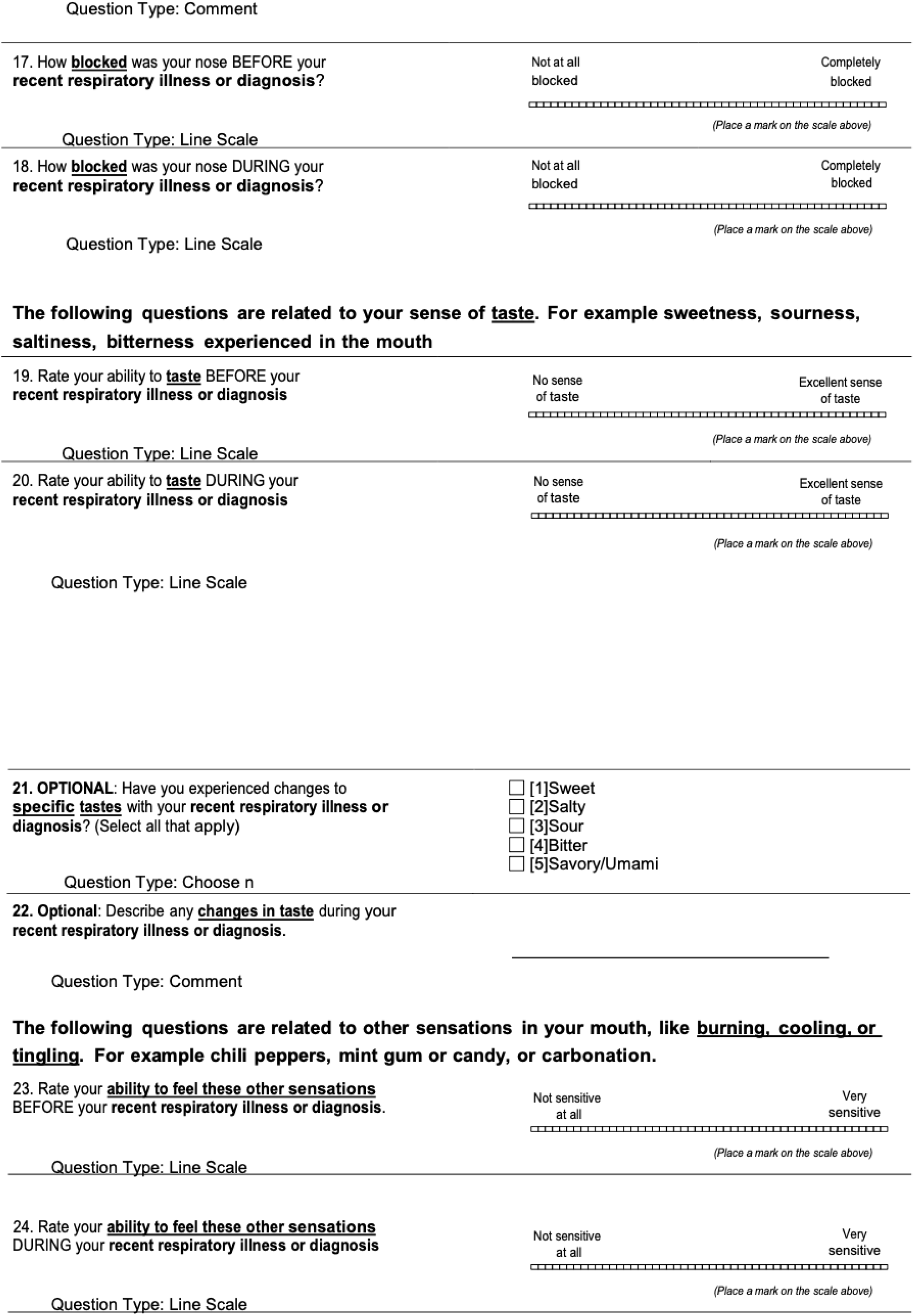

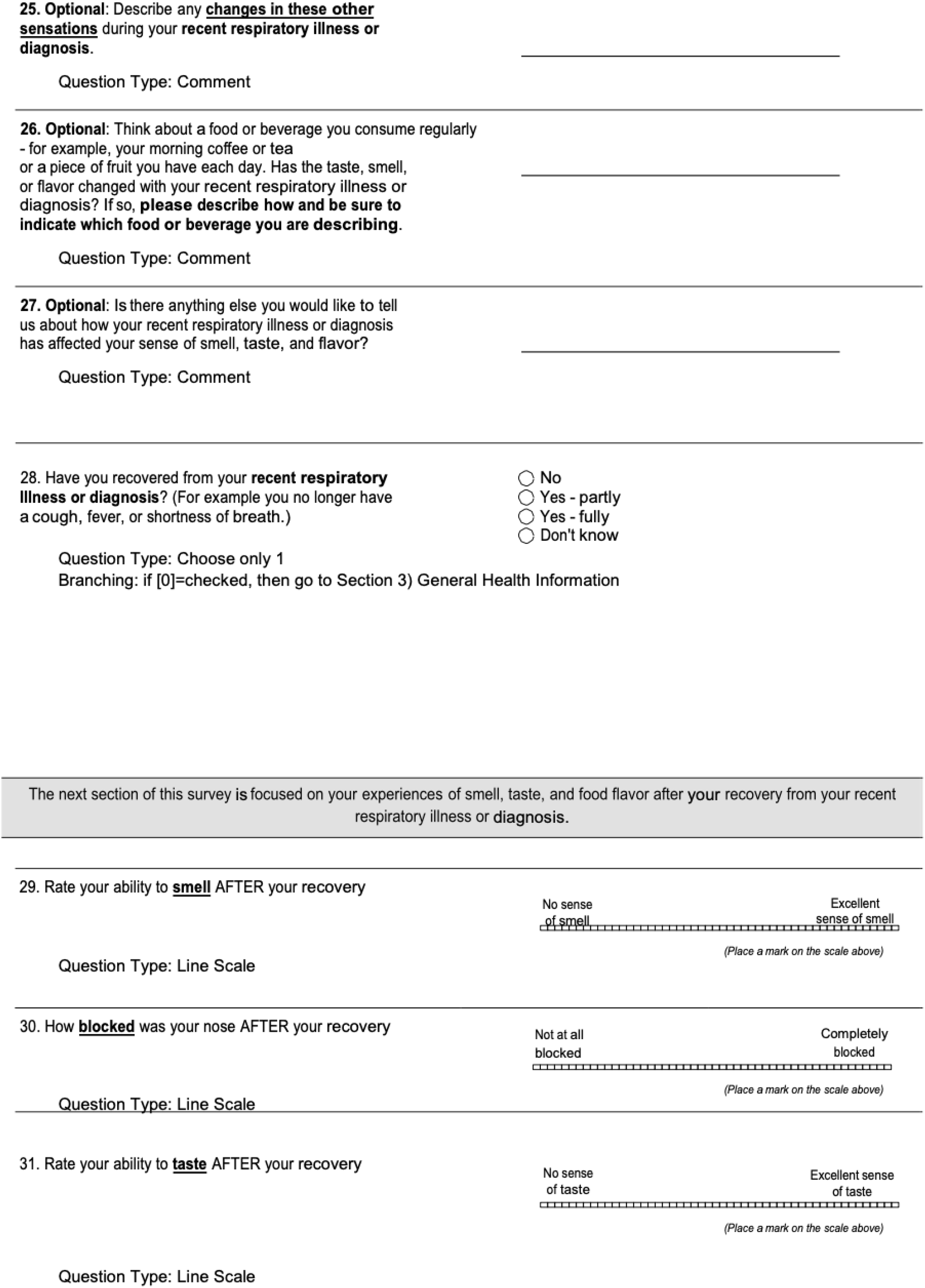

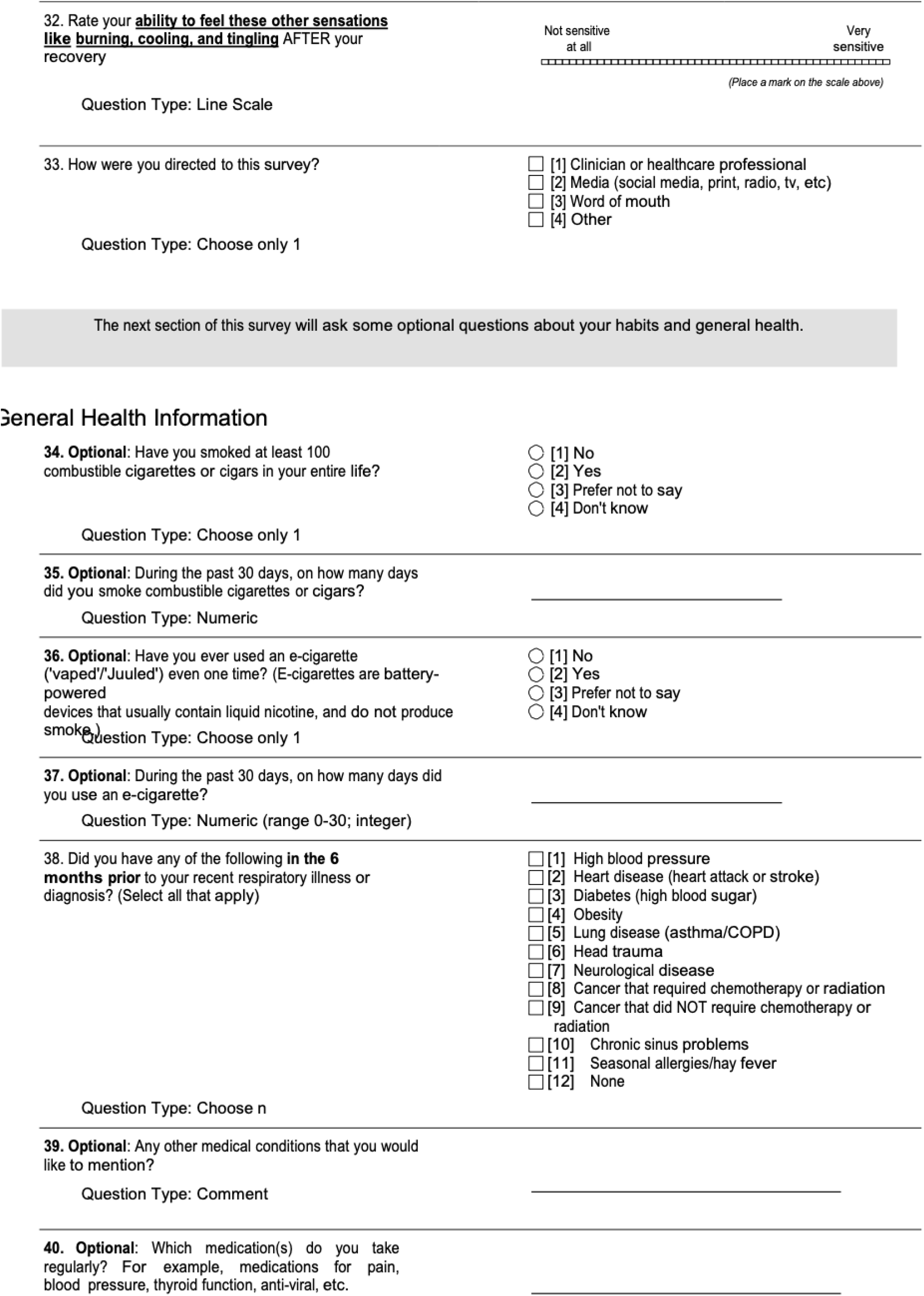

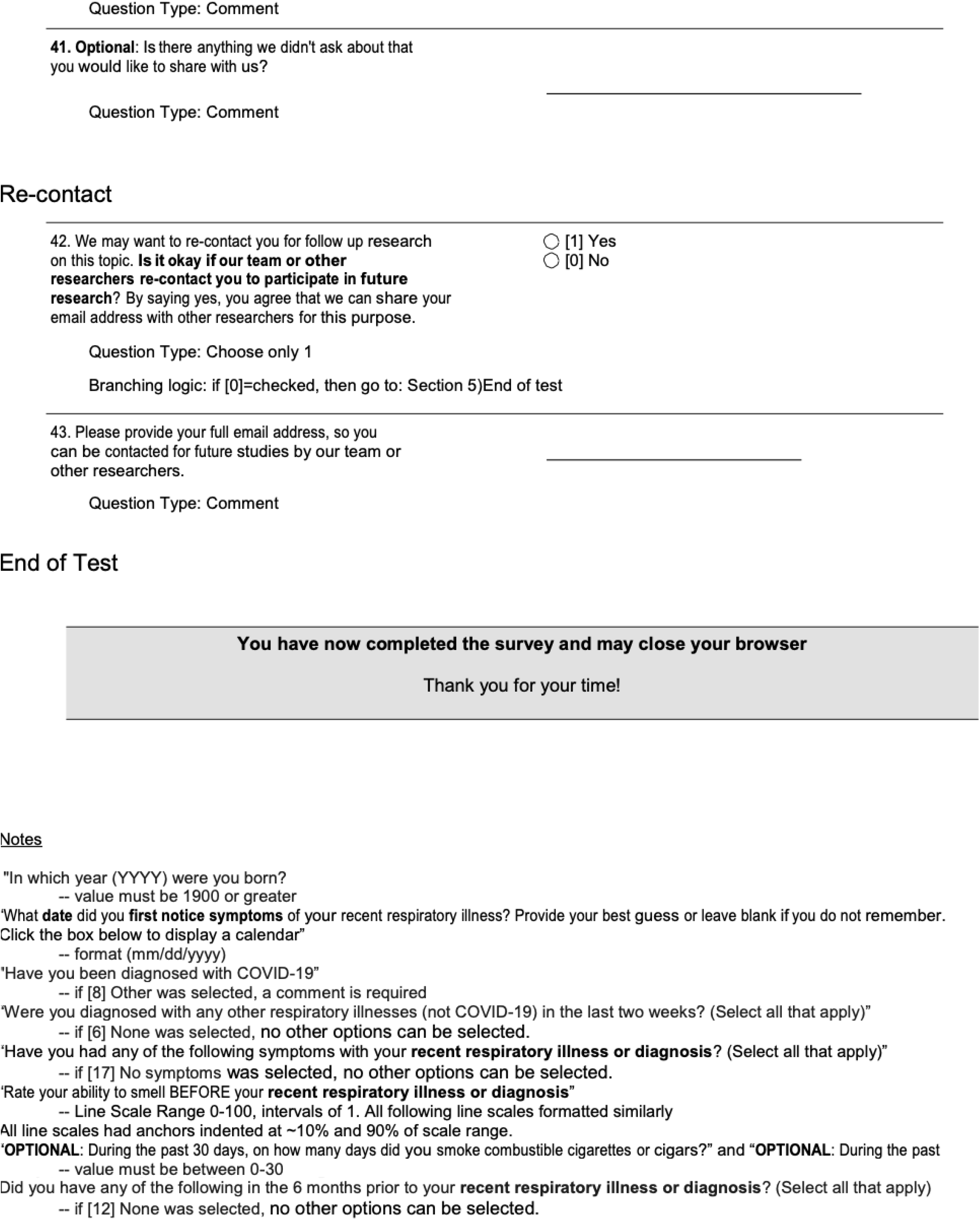

